# Controlled human malaria infection with PvW1 – a new clone of *Plasmodium vivax* with high quality genome assembly

**DOI:** 10.1101/2021.07.23.21259839

**Authors:** Angela M. Minassian, Yrene Themistocleous, Sarah E. Silk, Jordan R. Barrett, Alison Kemp, Doris Quinkert, Carolyn M. Nielsen, Nick J. Edwards, Thomas A. Rawlinson, Fernando Ramos Lopez, Wanlapa Roobsoong, Katherine J. Ellis, Jee-Sun Cho, Eerik Aunin, Thomas D. Otto, Adam J. Reid, Florian Bach, Geneviève M. Labbé, Ian D. Poulton, Arianna Marini, Marija Zaric, Margaux Mulatier, Raquel Lopez Ramon, Megan Baker, Celia H. Mitton, Jason C. Sousa, Nattawan Rachaphaew, Chalermpon Kumpitak, Nongnuj Maneechai, Chayanut Suansomjit, Tianrat Piteekan, Mimi M. Hou, Baktash Khozoee, David J. Roberts, Alison M. Lawrie, Andrew M. Blagborough, Fay L. Nugent, Iona J. Taylor, Kimberly J. Johnson, Philip J. Spence, Jetsumon Sattabongkot, Sumi Biswas, Julian C. Rayner, Simon J. Draper

**Author notes:** These authors contributed equally (Angela M. Minassian, Yrene Themistocleous). Corresponding authors: AMM; SJD.

## Abstract

Controlled human malaria infection (CHMI) provides a highly informative means to investigate host-pathogen interactions and enable *in vivo* proof-of-concept efficacy testing of new drugs and vaccines. However, unlike *Plasmodium falciparum*, well-characterized *P. vivax* parasites that are safe and suitable for use in modern CHMI models are limited. Here, two healthy malaria-naïve UK adults with universal donor blood group were safely infected with a clone of *P. vivax* from Thailand by mosquito-bite CHMI. Parasitemia developed in both volunteers and, prior to treatment, each volunteer donated blood to produce a cryopreserved stabilate of infected red blood cells. Following stringent safety screening, the parasite stabilate from one of these donors (“PvW1”) was thawed and used to inoculate six healthy malaria-naïve UK adults by blood-stage CHMI, at three different dilutions. Parasitemia developed in all volunteers, who were then successfully drug treated. PvW1 parasite DNA was isolated and sequenced to produce a high quality genome assembly by using a hybrid assembly method. We analysed leading vaccine candidate antigens and multigene families, including the Vivax interspersed repeat (VIR) genes of which we identified 1145 in the PvW1 genome. Our genomic analysis will guide future assessment of candidate vaccines and drugs, as well as experimental medicine studies.

## Introduction

The majority of human malaria is caused by two species of parasite – *Plasmodium falciparum* and *P. vivax*. Infection is initiated by an infected *Anopheles* mosquito bite, delivering sporozoites which rapidly migrate to and infect the liver. Asexual replication in the liver sees each infected cell produce thousands of merozoites. These rupture out into the blood and infect red blood cells (RBC), before undergoing exponential growth that leads to clinical symptoms and the associated morbidity and mortality. *P. vivax* is the predominant cause of malaria outside of Africa and is more geographically widespread than *P. falciparum*, with 2.5 billion people living at risk in Latin America, Oceania, Asia and the horn of Africa (1). Moreover, recent data demonstrate a significant burden of morbidity and associated mortality in young children and pregnant women (2), challenging the long-held dogma that this parasite is “benign” (3).

A number of factors also underlie the differing epidemiology of *P. vivax* and make it more difficult to control and eliminate than *P. falciparum* (4). Most notably earlier development of gametocytes leads to transmission prior to symptom onset, and its ability to form dormant liver-stage forms, termed “hypnozoites”, causes waves of relapsing bood-stage parasitemia and sustained transmission (5). However, despite a clear global health need to develop an effective vaccine and improved antimalarial drugs, these efforts continue to lag behind those for *P. falciparum*. The reasons for this are numerous, but perhaps most significant is the fact that *P. vivax* has not been able to be adapted to long-term *in vitro* culture, despite extensive efforts. This has severely limited laboratory studies, as well as the development of modern controlled human malaria infection (CHMI) models, which rely on a well-defined isolate of *P. vivax*, and would enable *in vivo* efficacy testing of candidate vaccines and anti-malarial drugs in proof-of-concept clinical trials. This is in contrast to *P. falciparum* where *in vitro* culture and sophisticated genetic modification experiments are carried out all over the world, and CHMI can be initiated by the traditional mosquito-bite method, or by injection of cryopreserved sporozoites or an inoculum of blood-stage parasites (6). Most of these studies have been carried out in non-endemic settings, but CHMI trial capacity is now expanding across endemic countries in Africa, enabled by the use of cryopreserved sporozoites. In contrast, modern CHMI with *P. vivax* has been less utilized, with only a handful of studies reported (7).

For mosquito-bite *P. vivax* CHMI trials, most have taken place in Cali, Columbia (8–11) plus one at the Walter Reed Army Institute of Research (WRAIR), USA (12), with 108 volunteers challenged in total. Such trials necessitate production of infected mosquitoes in an endemic setting using fresh gametocytes from an infected patient. Shipment of the mosquitoes to non-endemic areas, and timing these activities with recruitment of volunteers who may receive an intervention such as a vaccine, poses significant logistical challenges. Moreover, a different isolate of *P. vivax* is inevitably used for every trial which can hamper interpretation of the results and inter-study comparability. These studies also pose the risk of relapse, and thus require participants to be screened for glucose-6-phosphate dehydrogenase (G6PD) deficiency (to avoid hemolysis induced by primaquine treatment). They also require assessment of the volunteers’ ability to metabolize primaquine, given relapsing infection occurred in two volunteers in the CHMI study at WRAIR despite primaquine treatment. Here, drug failure was subsequently linked to the volunteers’ cytochrome P450 2D6 (CYP2D6) genotypes that were predicted to be poor or intermediate metabolizer phenotypes of the drug (13).

The use of the blood-stage CHMI model (14, 15) has several advantages over mosquito-bite CHMI, although does not mimic the route of natural infection. Here a cryopreserved stabilate of infected RBC (iRBC) is produced from a donor volunteer, enabling subsequent direct blood-stage inoculation of other volunteers with small numbers of parasites. This model is more practical in non-endemic settings; enables access to the parasite’s genetic data before CHMI; removes all risk of relapsing infection; and enables multiple studies with the same strain of parasite (for which a safety database can be established). In the case of *P. falciparum*, this model has also proved particularly suitable for estimating the blood-stage parasite multiplication rate (PMR) (16) and for enabling experimental transmission to mosquitoes (17), as compared to studies initiated by mosquito-bite. The blood-stage model is also advantageous because it extends the period of blood-stage infection, allowing for longer studies of the human immune response and also switching/selection of parasite variant surface antigens (18).

Two cryopreserved stabilates of blood-stage *P. vivax* have been reported to-date, both produced by the group at the QIMR Berghofer Medical Research Institute, Australia and obtained from returning travellers who donated infected blood prior to treatment. The first isolate, HMPBS-*Pv* from the Solomon Islands, was safely tested by blood-stage CHMI in eight volunteers (19, 20), however this necessitated recruitment of individuals with blood group A to match that of the donor. The second *P. vivax* isolate, HMP013-*Pv*, was from India and a blood group O+ donor. This has been tested in healthy adult volunteers and showed successful induction of gametocytemia and experimental transmission of *P. vivax* from humans to mosquitoes (21), and also enabled trials of candidate drugs and further methodology development (22, 23).

Here we take a significant step forward for *P. vivax* CHMI by establishing a well-characterized Thai clone of *P. vivax* suitable for both mosquito-bite and blood-stage CHMI. We elected to produce a cryopreserved stabilate of iRBC from blood donated by healthy volunteers infected via mosquito-bite CHMI, as opposed to using a blood donation from a returning traveller. This provided numerous advantages in terms of logistical timing, and our ability to recruit in advance volunteers who passed a full health screen and who had universal donor blood group. In real-time we were able to select mosquitoes infected in Thailand with a single *P. vivax* genotype, thus avoiding production of a cryopreserved iRBC stabilate from a polyclonal infection. It also minimized the time from mosquito to blood bank (compared to infected returning travellers); this is important as it has previously been shown that mosquitoes reset parasite virulence and expression of variant surface antigens (24). Following production of the cryopreserved parasite stabilate, which we called “PvW1”, we demonstrated safety and infectivity by blood-stage CHMI in six healthy adults, and we also report a full genomic analysis of the new PvW1 clone.

## Results

### Source patient case-finding and preparation of infected mosquitoes

For infection of mosquitoes, source patients were recruited from a medical clinic in southern Thailand. Patient blood samples that tested positive for *P. vivax* and negative for filarial disease were fed to *Anopheles dirus* mosquitoes via a direct membrane feeding system in Thailand. Oocyst and sporozoite counts subsequently confirmed successful production of three independent batches of infected mosquitoes (**Fig. S1A**). In parallel, and in real-time, source patient samples underwent additional and rigorous testing in the UK for blood-borne infections and mosquito-borne diseases other than malaria; all tests were negative. Nested PCR reported mono-infection with *P. vivax* (**Fig. S1B,C**), thus confirming the diagnosis in Thailand, however, genotyping analysis suggested that only one blood sample (C05-001) contained a single *P. vivax* genotype (**Fig. S1A**). Mosquitoes fed off this patient’s blood were therefore selected and shipped from Thailand to the UK.

### Screening of healthy UK volunteers for blood donation

In parallel we enrolled two healthy UK adult volunteers into the VAC068 clinical trial (**Fig. S2**). These volunteers were specifically screened to be universal blood donors (blood group O rhesus-negative), Duffy-blood group positive (7, 25); G6PD normal (26); and to have a CYP2D6 genotype predicted to be an extensive metabolizer phenotype (27) alongside satisfactory demonstration of primaquine metabolism following administration of a single test dose of drug (13) (**Table S1**, **Fig. S3**). Each volunteer also underwent an extensive screen for blood-borne infections; all test results were negative, except both participants were IgG seropositive for Epstein-Barr virus (EBV) and cytomegalovirus (CMV) (**Table S1**), indicating past infection. However, we did not exclude volunteers based on their serostatus for these two viruses.

### Mosquito-bite CHMI – safety and parasite growth dynamics

For the C05-001 mosquito batch, the mean [range] number of oocysts per mosquito was 3 [0-6] at day 7 post-feeding and the median score for number of sporozoites observed in the salivary glands at day 14 post-feeding was +2 (defined as >10-100 sporozoites) (**Fig. S1A**). This was relatively low but agreed to be sufficient for human transmission. Subsequently, the two healthy UK adult volunteers screened and consented to take part in VAC068 were each exposed to five “infectious bites” as defined post-skin feeding by microscopic examination of each mosquito. To achieve this, volunteers 01-004 and 01-008 required 17 and 33 mosquitoes, respectively, to bite their arm.

Parasites were first reliably detected in the blood of both volunteers by qPCR at the evening clinic visit 8 days post-CHMI (dC+8.5), and parasitemia then steadily rose over time (**Fig. 1A**, **Table S2**). Over the course of the CHMI period, the two volunteers experienced a range of solicited adverse events (AEs), with both reporting grade 3 fatigue and at least grade 2 anorexia, chills, feverishness, headache, malaise, nausea and sweats (**Fig. 1B**). Both volunteers were admitted for blood donation when they met protocol-specified criteria defined by symptoms and or threshold levels of parasitemia as measured in genome copies (gc)/mL by qPCR. This occurred on the morning of dC+14 for both volunteers, who both crossed the 10,000 gc/mL threshold on dC+13.5 and developed fever on dC+14. Following admission to the clinical trials unit, a 250 mL blood sample was collected (at dC+14 for volunteer 01-008 and dC+14.5 for volunteer 01-004); both were positive by thick film microscopy and reported 16,717 or 31,010 gc/mL by qPCR, respectively. Prior to cryopreservation, these blood samples were then randomized and relabelled either “Donor 1” or “Donor 2” and are now referred to as such in the Results.

**Figure 1.**
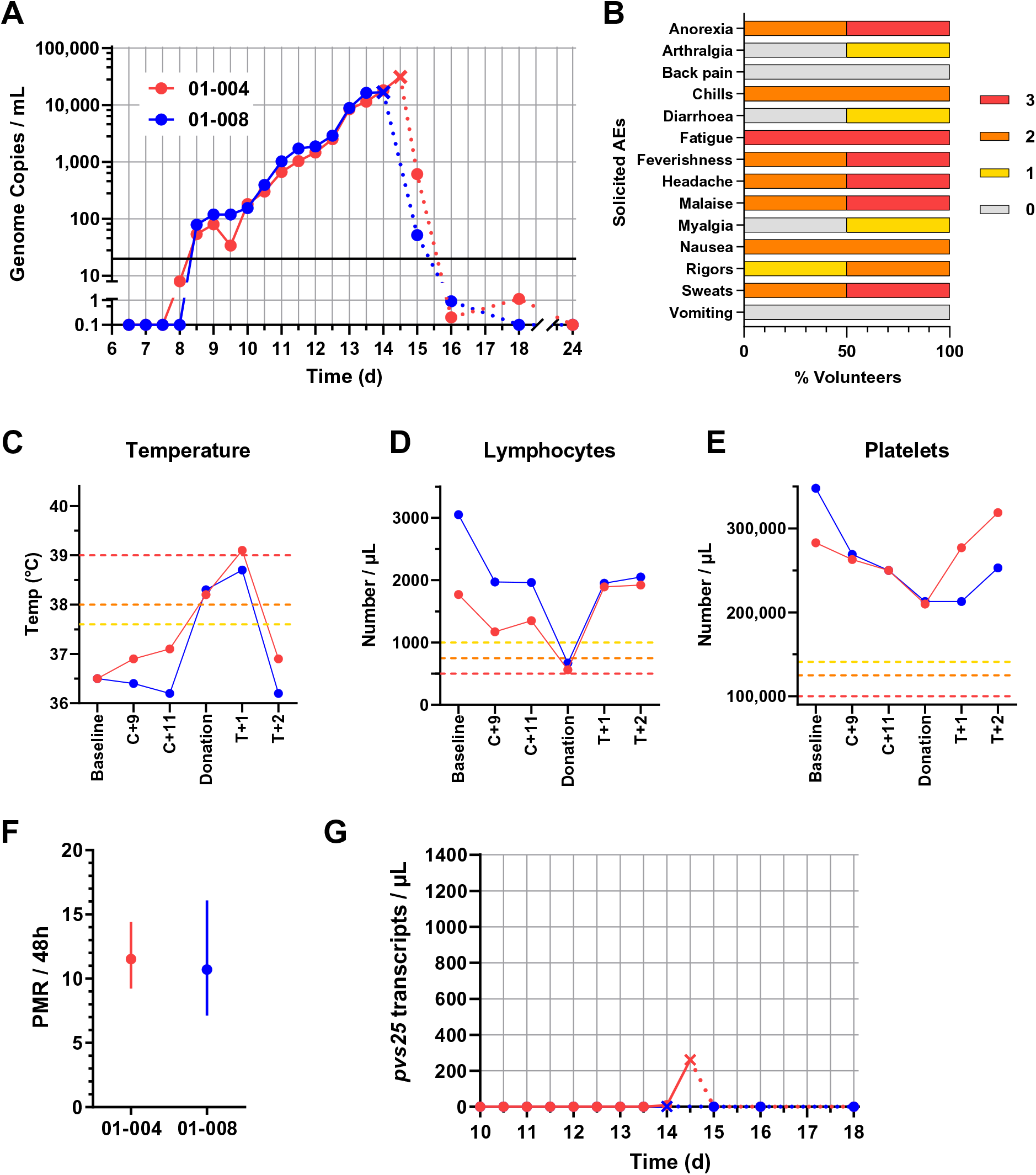
Safety and parasite growth dynamics of *P. vivax* sporozoite CHMI. (**A**) qPCR data for the VAC068 trial. Parasitemia measured in genome copies / mL is shown over time for each volunteer. CHMI was initiated by mosquito bite on day 0. Cross symbols indicate the time-point of blood donation followed by anti-malarial treatment. Solid lines show qPCR readouts pre-treatment, and dotted lines post-treatment. Solid black line indicates 20 gc/mL (the minimum level to meet positive reporting criteria); samples below this are shown for information only. (**B**) The solicited systemic adverse events (AEs) recorded during the CHMI period (from 1 day up until 45 days post-challenge) are shown as the maximum severity reported by each volunteer and as a percentage of the volunteers reporting each individual AE (n=2). Color-coding refers to AE grading: 0 = none; 1 = mild; 2 = moderate; 3 = severe. (**C**) Volunteer temperature (maximum self-recorded by volunteer or measured in clinic) at the indicated time-points: baseline pre-CHMI; 9 and 11 days post-CHMI (C+9, C+11); time of blood donation; and 1 and 2 days post-treatment (T+1, T+2). AE grading cut-offs are indicated by the dotted lines (yellow = grade 1; orange = grade 2; red = grade 3). (**D**) Lymphocyte and (**E**) platelet counts plotted as for panel C. (**F**) The PMR per 48 h was modelled from the qPCR data up until the time-point of blood donation/treatment; PMR ± 95% CI is shown for each volunteer. (**H**) Gametocytemia was assessed over time by qRT-PCR for *pvs25* transcripts; symbols and lines as per panel A.

After blood donation, each volunteer was immediately treated with Riamet® followed by a 14-day course of primaquine; no supportive treatment or hospital admission was required for either volunteer. Monitoring by qPCR on days 1, 2, 4, 10 and 16 post-treatment showed a rapid decline in blood-stage parasitemia followed by negative readings for both volunteers (**Fig. 1A**, **Table S2**). Most solicited symptoms increased in severity in the first 24 hours after starting anti-malarial treatment (**Fig. S4A**). Objective fever also increased in the 24 hours post-treatment (**Fig. 1C**), and one volunteer developed a grade 3 pyrexia (**Fig. S4B**), but all symptoms had completely resolved within 5 days of starting treatment. Both volunteers also experienced some short-lived grade 1 or 2 AEs possibly related to the anti-malarial treatment (dizziness, insomnia and abdominal pain) (**Fig. S4C**). Very few unsolicited AEs (at least possibly related to CHMI) were reported by either volunteer (**Table S3A**), and only one grade 3 unsolicited AE (migraine, not related to CHMI) was reported by 01-004 more than 2 months post-challenge, requiring attendance to their doctor and resolving within 48 hours (**Table S3B**). Lymphocyte and platelet counts dropped in both volunteers around the time of blood donation (platelets remained within the normal range but both developed a grade 2 lymphocytopenia), rising back to pre-challenge levels within 48 hours (**Fig. 1D-E** and **Table S3C**). Volunteer 01-008 also developed a transient grade 1 anemia ∼6 weeks post-challenge (123 g/L at dC+47) which may or may not have been related to CHMI, but this resolved within 3 months (131 g/L at dC+94).

Following completion of the study, the parasite multiplication rate (PMR) for both volunteers was calculated using a linear model fitted to log_10_-transformed qPCR data (28). These data showed comparable PMRs in both volunteers, with 10.7- and 11.5-fold growth per 48 hours (**Fig. 1G**). We also analysed gametocytemia using a qRT-PCR assay to detect mature female gametocyte *pvs25* transcripts. Volunteer 01-004 showed only low levels at the final time-point pre-treatment (dC+14.5) whilst none were detected in volunteer 01-008 (**Fig. 1H**).

Finally, with regard to longer-term safety monitoring, clinic visits at dC+45 and dC+90 gave rise to no safety concerns or indication of relapsing infection, and repeat serological tests for blood-borne infections at dC+90 all remained negative. Ongoing annual follow-up by email will continue for 5 years post-CHMI, however, as of time of writing (3 years post-primaquine treatment) no relapse of *P. vivax* has been diagnosed for either volunteer (see **Supplementary Text**).

### Cryopreservation and *in vitro* testing of *P. vivax* infected blood

After blood donation, the leukodepleted blood from both volunteers in VAC068 was processed, and the RBC mixed with Glycerolyte 57 to form a stabilate prior to cryopreservation. In total, 190 vials were frozen for Donor 1 and 185 for Donor 2. In process testing by qPCR indicated minimal or no loss of parasites during filtration (95% and 105% recovery for Donor 1 and Donor 2, respectively). We next tested for parasite viability in both cryopreserved stabilates. Vials were thawed and cells used in a short-term *in vitro* parasite culture assay, given *P. vivax* cannot currently be cultured long-term *in vitro*. Parasite growth was detectable by qPCR and light microscopy through one initial growth cycle in samples collected from Donor 1, with normal progression of parasite morphology seen on Giemsa stained thick and thin blood films (**Fig. 2**). However, no growth was discernible in samples obtained from Donor 2. We therefore undertook further QC testing on vials from Donor 1, with the material tested for sterility, mycoplasma and endotoxin; all tests were passed. Another screen for blood-borne infections was also conducted on the plasma derived directly from the blood donation; all tests were negative.

**Figure 2.**
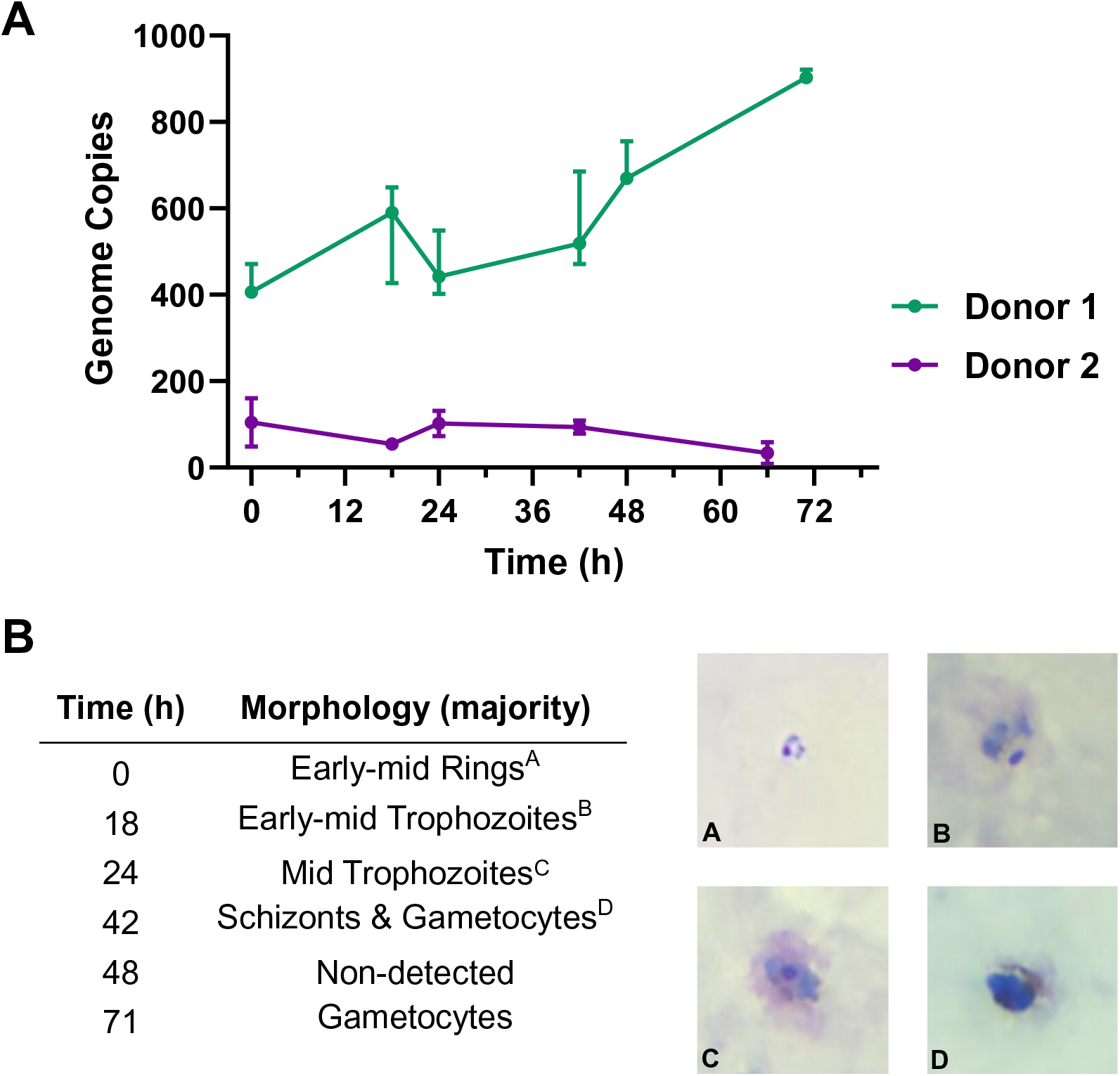
Test of cryopreserved parasite viability by short-term *in vitro* culture assay. (**A**) Test vials of cryopreserved parasites from Donor 1 and Donor 2 were thawed and cells used in a short-term *in vitro* parasite culture assay. *P. vivax* parasite growth was monitored by qPCR in 20 µL samples of RBC extracted at the indicated time-points. Median and range of triplicate readings are shown in genome copies measured per 20 µL sample. (**B**) Parasite morphology was monitored at the same time-points over the first growth cycle by light microscopy of Giemsa-stained thick and thin blood films Representative images are shown from Donor 1, and the predominant morphology observed is reported.

Finally, we also screened Donor 1 for the Kell blood group antigen because women of childbearing potential that receive a blood transfusion have a small additional risk of developing RBC alloantibodies that could cause problems during pregnancy. In particular, there is a potential risk of development of hemolytic disease of the newborn in relation to Kell antigen incompatibility, i.e. if Kell-positive donor blood is transfused to a Kell-negative female recipient. However, testing of the donor’s blood sample confirmed Kell antigen negativity, thereby allowing future universal administration of the cryopreserved *P. vivax* iRBC stabilate with respect to gender.

### Blood-stage CHMI – PvW1 infectivity, parasite growth dynamics and safety

Given all safety and viability tests were passed for the cryopreserved stabilate of *P. vivax* iRBC from Donor 1, we named this clonal isolate “PvW1” and proceeded to test safety and infectivity by blood-stage CHMI. We therefore recruited six healthy, malaria-naïve UK adults into the VAC069A clinical trial, comprising three groups of two volunteers (**Fig. S5**), and tested feasibility of infection at three different doses of PvW1 blood-stage inoculum. Five vials of the PvW1 cryopreserved stabilate were thawed and then combined to produce a single batch of blood-stage inoculum. Two volunteers receive a whole vial’s worth of iRBC (“neat”), two volunteers received one fifth of the challenge dose via a 1:5 dilution, and the final two volunteers were inoculated with one twentieth of the dose via a 1:20 dilution. All six volunteers underwent blood-stage CHMI at the same time.

Blood-stage parasitemia was monitored as previously by qPCR, beginning one day after challenge (dC+1) (**Fig. 3A**, **Table S4**). All six volunteers were successfully infected, with a median time to diagnosis of 15.25 days post-CHMI (range 12.5-16.5) (**Fig. 3B**). The median (range) parasitemia at diagnosis across all six volunteers was 9,178 (3,779 – 17,795) gc/mL (**Fig. 3C**). We also calculated the PMR as before using a linear model fitted to log_10_-transformed qPCR data (28). These data showed a median (range) of 5.7 (3.6 – 7.0)-fold growth per 48 hours across the six volunteers (**Fig. 3D**), notably lower than that previously observed in the mosquito-bite CHMI study (**Fig. 1F**). There was also no discernible difference in the PMRs across the three different challenge dose cohorts (**Fig. 3D**); or across the three different Duffy blood group sero-phenotypes, all with median values between 5.3 – 5.9-fold growth per 48 hours (**Fig. 3E**). We also analysed gametocytemia at the six time-points preceding diagnosis for each volunteer, and observed rising levels in all individuals (**Fig. 3F**). This was in clear contrast to the observations post-mosquito bite CHMI (**Fig. 1G**) and despite comparable (if not slightly lower) levels of overall blood-stage parasitemia as measured in gc/mL. Here, we also saw a strong positive correlation between the measured overall levels of parasitemia in gc/mL versus *pvs25* transcripts/µL (**Fig. 3G**).

**Figure 3.**
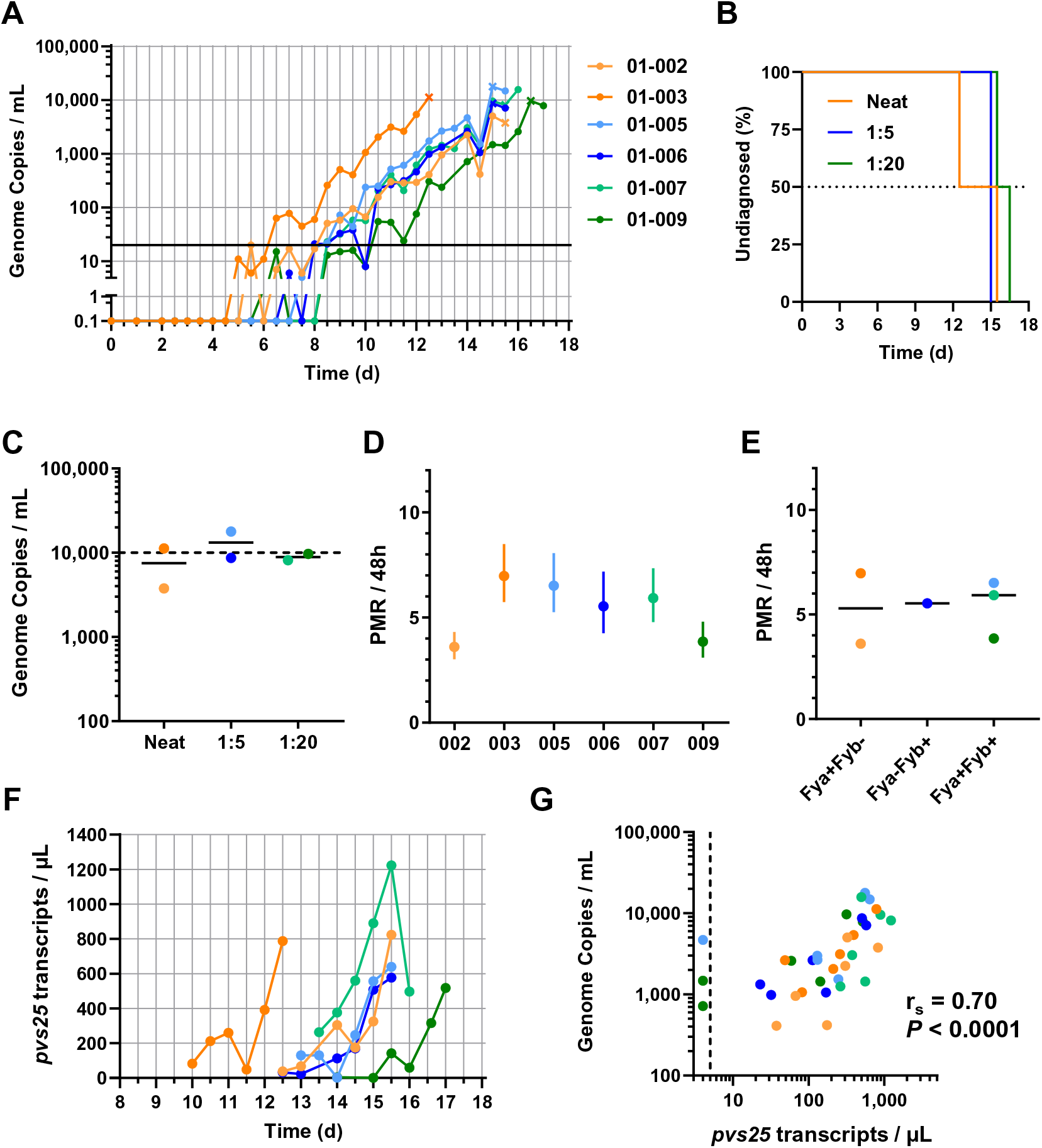
Parasite growth dynamics of *P. vivax* PvW1 clone blood-stage CHMI. (**A**) qPCR data for the VAC069A trial. Parasitemia measured in genome copies (gc) / mL is shown over time for each volunteer. CHMI was initiated by blood-stage inoculation on day 0. Cross symbols indicate the time-point of diagnosis. Orange = neat inoculum dose; blue = 1:5 and green = 1:20 dilution of the neat inoculum dose. Solid black line indicates 20 gc/mL (the minimum level to meet positive reporting criteria); samples below this are shown for information only. (**B**) Kaplan-Meier plot of time to diagnosis in days for the VAC069A study. (**C**) Parasitemia measured in gc/mL at the time-point of diagnosis. Individual data and median are indicated for each dose group. (**D**) The PMR per 48 h was modelled from the qPCR data up until the time-point of diagnosis; PMR ± 95% CI is shown for each volunteer. (**E**) Individual and median PMR are shown with volunteers grouped according to their Duffy blood group antigen (Fy) serological phenotype. (**F**) Gametocytemia was assessed over time by qRT-PCR for *pvs25* transcripts; colored lines as per panel A. (**G**) Correlation of total parasitemia measured in gc/mL versus *pvs25* transcripts/µL. Spearman’s rank correlation coefficient and *P* value are shown, n=36.

With regards to safety, there were no serious adverse events (SAEs) in the VAC069A study and all volunteers completed treatment without complication. One volunteer withdrew at dC+28, with the remaining five completing clinical follow-up at dC+90 (**Fig. S5**). The maximum severity of solicited AEs at any time during the CHMI period is shown for all six volunteers in **Fig. 4A**, with four volunteers reporting grade 3 solicited AEs (most commonly feverishness) persisting for 24 hours and one for 48 hours (**Table S5A**). The proportion of volunteers reporting solicited AEs specifically pre-diagnosis, peri-diagnosis and post-treatment is shown in **Fig. 4B**. Around the time of diagnosis, 33-50% of the volunteers reported mild-to-moderate symptoms; mainly fatigue, headache, myalgia, malaise, feverishness and chills. Symptoms peaked in severity in the first 24 hours after starting anti-malarial treatment with Riamet® or Malarone, with only one volunteer remaining asymptomatic (**Fig. 4B**). Objective fever also increased in the 24 hours post-treatment, with 3/6 volunteers developing pyrexia, one of each grades 1-3 (**Fig. 4C**, **Fig. S6A**). Nevertheless, most symptoms had completely resolved within a few days of starting treatment and only one volunteer still had headache and fatigue at 6 days post-starting treatment (T+6) (**Fig. 4B**). Three volunteers (50%) also experienced short-lived AEs possibly related to the anti-malarial drugs (50% moderate dizziness, 33% mild insomnia, cough and palpitations) (**Fig. S6B**). Very few unsolicited AEs (at least possibly related to CHMI) were reported by any of the volunteers (**Table S5B**).

**Figure 4.**
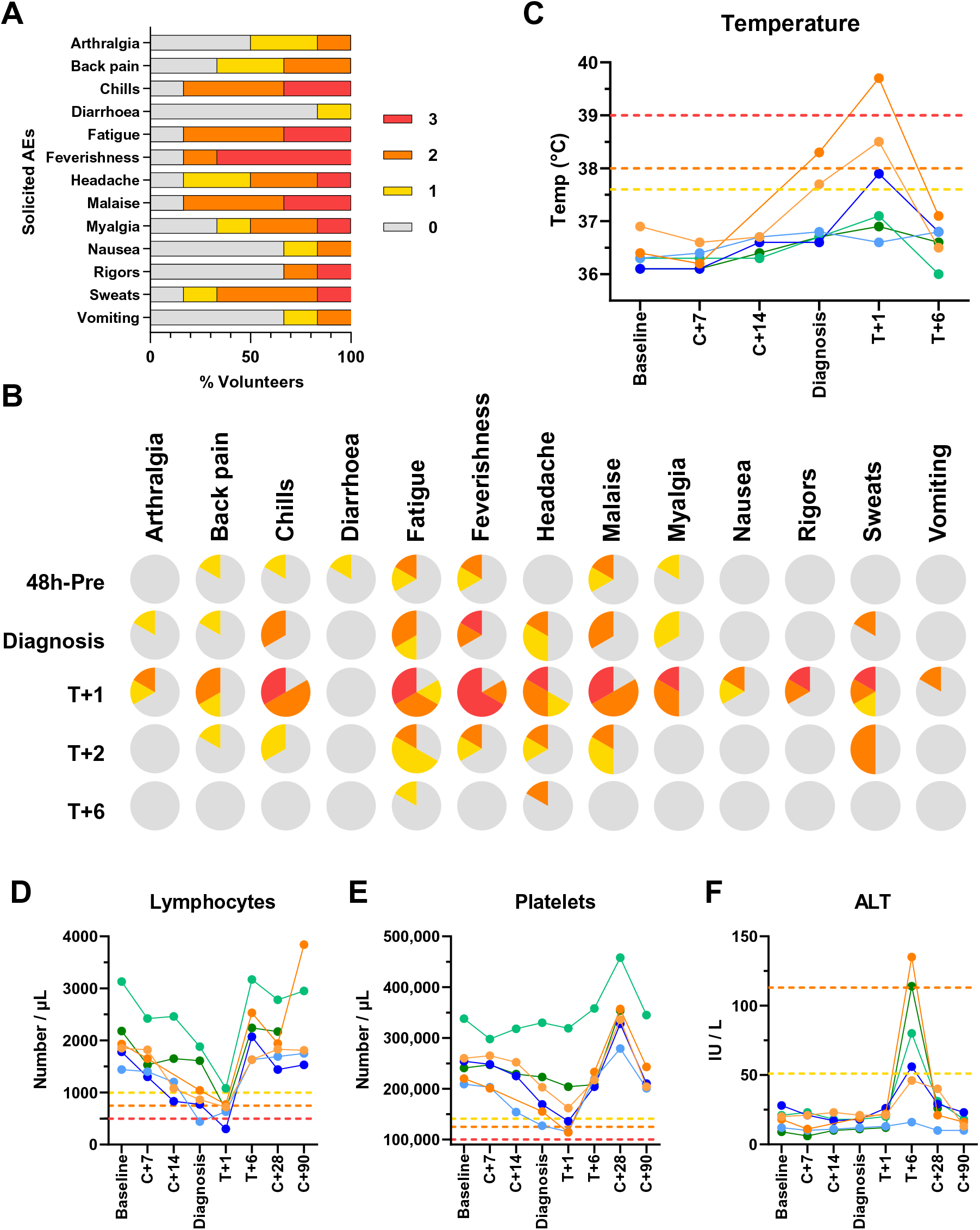
Safety analysis of *P. vivax* PvW1 clone blood-stage CHMI. (**A**) The solicited systemic adverse events (AEs) recorded during the CHMI period (from 1 day up until 90 days post-challenge) are shown as the maximum severity reported by each volunteer and as a percentage of the volunteers reporting each individual AE (n=6). Color-coding refers to AE grading: 0 = none; 1 = mild; 2 = moderate; 3 = severe. (**B**) The solicited systemic AEs recorded at the indicated time-points during the CHMI period are shown as the maximum severity reported by each volunteer and as a percentage of the volunteers reporting each individual AE (n=6). Color-coding as per panel A. 48h-pre = the 48 hour period prior to *P. vivax* diagnosis; Diagnosis = time-point of diagnosis; +1, +2 and +6 days post-treatment (T). (**C**) Volunteer temperature (maximum self-recorded by volunteer or measured in clinic) at the indicated time-points: baseline pre-CHMI; 7 and 14 days post-CHMI (C+7, C+14); time of diagnosis; and 1 and 6 days post-treatment (T+1, T+6). AE grading cut-offs are indicated by the dotted lines (yellow = grade 1; orange = grade 2; red = grade 3). (**D**) Lymphocyte and (**E**) platelet counts, and (**F**) alanine aminotransferase (ALT) measurements, all plotted as for panel C but also including C+28 and C+90 time-points.

With regard to laboratory AEs (**Table S5C**), lymphocyte counts dropped significantly in 4/6 volunteers around the time of diagnosis or 1 day post-treatment (grade 3 lymphocytopenia in two volunteers), but all counts normalized within 6 days of starting treatment (**Fig. 4D**). Two volunteers developed a short-lived grade 2 thrombocytopenia, again normalizing within 6 days of treatment (**Fig. 4E**); whilst two volunteers also developed a mild-moderate anaemia post-diagnosis. With regards to the latter, one normalized within 28 days of challenge, the other persisted at grade 1 at dC+90 (102 g/L) and so was referred to their medical practitioner for ongoing monitoring as a precautionary measure (**Table S5C** and **Fig. S6C**). The only notable change in blood chemistry was a transient grade 1-2 rise in the ALT in 4/6 volunteers, captured consistently at 6 days post-treatment (**Fig. 4F**). All fully resolved to pre-challenge levels with no associated abnormalities in other indices of liver function (**Fig. S6C**, **Table S5D**). Finally, we also confirmed CMV and EBV sero-status of all volunteers pre- and post-CHMI. All six volunteers were EBV sero-positive pre-CHMI and three were CMV sero-positive. Of the three CMV sero-negative volunteers, one withdrew consent and left the trial at C+28 and was therefore not re-tested, whilst the other two remained sero-negative when re-tested at C+90.

### Antibody responses to blood-stage merozoite antigens post-CHMI

We next assessed for the induction of serum IgG antibody responses post-CHMI against two well-known blood-stage merozoite antigens – *P. vivax* merozoite surface protein 1 C-terminal 19 kDa region (PvMSP1_19_) and *P. vivax* Duffy-binding protein region II (PvDBP_RII). All volunteers had detectable IgG against PvMSP1_19_ post-CHMI, with similar results seen in the VAC068 mosquito-bite sporozoite CHMI study and the VAC069A blood-stage CHMI study (**Fig. 5A**). However, there were no detectable responses post-CHMI against PvDBP_RII in any of the volunteers, in contrast to positive control samples from a cohort of healthy UK adult volunteers previously vaccinated with the PvDBP_RII antigen (29) which were included here for comparison (**Fig. 5B**).

**Figure 5.**
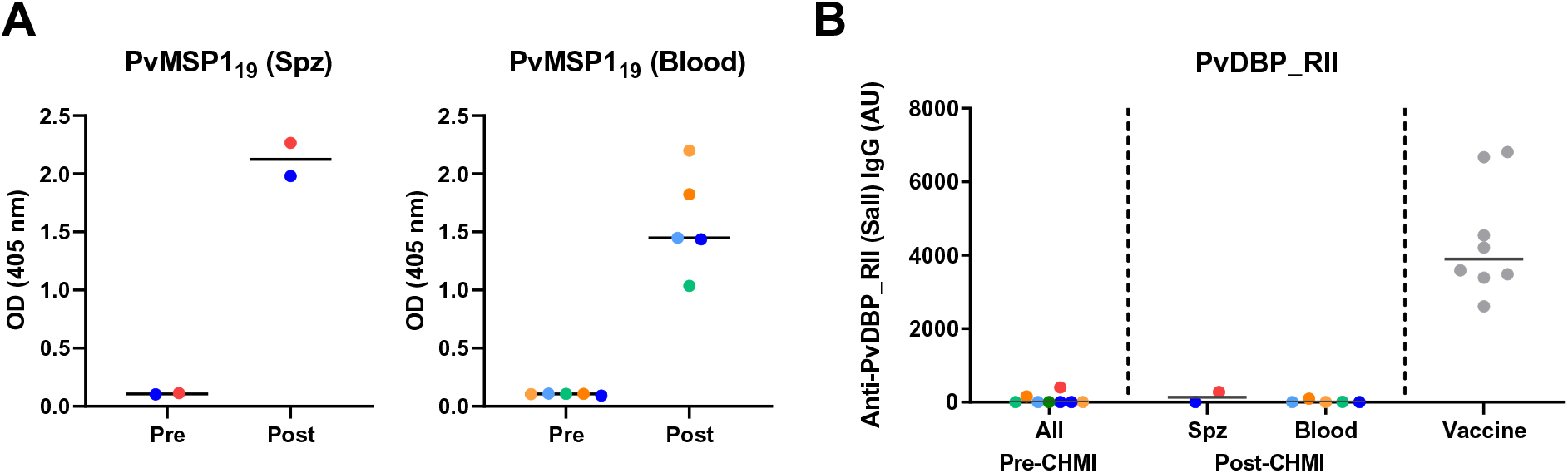
Induction of serum antibody responses to merozoite antigens during CHMI. (**A**) Serum anti-PvMSP1_19_ IgG ELISA was conducted on samples from the VAC068 mosquito-bite / sporozoite (spz) CHMI study (n=2) and the VAC069A blood-stage CHMI study (n=5 as one volunteer withdrew at dC+28). Optical density (OD) 405nm data are shown for sera tested at a 1:100 dilution from the pre-CHMI (dC-1) and 90 days post-CHMI (dC+90) time-points. Samples color-coded as per previous figures. (**B**) Serum anti-PvDBP_RII (SalI allele) IgG as measured by standardized ELISA, reporting in arbitrary units (AU). Same samples tested as in panel A. Vaccine = positive control samples (n=8) from a previous Phase Ia clinical trial of a PvDBP_RII vaccine (29). Individual data and median are shown.

### PvW1 genome assembly allows resolution of complex multi-gene families

Finally, we produced a genome assembly for PvW1 by using a hybrid assembly method which combined long PacBio reads with short Illumina reads. The PvW1 genome assembled into 14 scaffolds (the 14 *P. vivax* chromosomes), and is comparable in both assembly size and number of genes to the highest quality existing *P. vivax* assembly, PvP01 (30) (**Table 1**). The PvW1 assembly has fewer unassigned scaffolds than any other assembly, indicating the completeness of the assembled genome and the benefits of using a combination of long and short reads; note the PvP01, PvC01 and PvT01 were all assembled using Illumina data only (30), while the original reference, PvSalvador-1 (SalI), was created using capillary sequence data (31).

**Table 1.** Comparison of genome assembly statistics between PvW1 and other P. vivax assemblies. PvW1 genome assembly statistics were compared with the best available existing assemblies: PvP01, PvC01, PvT01 and SalI (30, 31). pir = *P. vivax Plasmodium* interspersed repeat, also known as VIR.

| Genome Features | PvW1 | PvP01 | PvC01 | PvT01 | Sall |
| --- | --- | --- | --- | --- | --- |
| **Nuclear genome** |  |  |  |  |  |
| Assembly size (Mb) | 28.9 | 29 | 30.2 | 28.9 | 26.8 |
| G + C content (%) | 39.9 | 39.8 | 39.2 | 39.7 | 42.3 |
| No. scaffolds assigned to chrom. | 14 | 14 | 14 | 14 | 30 |
| No. unassigned scaffolds | 3 | 226 | 529 | 359 | 2745 |
| No. genes | 6583 | 6642 | 6690 | 6464 | 5433 |
| No. pir (VIR) genes | 1145 | 1212 | 1061 | 867 | 346 |
| **Mitochondrial genome** |  |  |  |  |  |
| Assembly size (bp) | 5994 | 5989 | - | - | 5990 |
| G + C content (%) | 30.5 | 30.5 | - | - | 30.5 |
| **Apicoplast genome** |  |  |  |  |  |
| Assembly size (kb) | 34.5 | 29.6 | 27.6 | 6.6 | 5.1 |
| G + C content (%) | 14.4 | 13.3 | 12.7 | 19.7 | 17.1 |
| No. genes | 54 | 30 | 3 | 0 | 0 |

The high quality of the PvW1 assembly allowed us to identify 1145 Vivax interspersed repeat (VIR) genes within the genome, comparable in number to the PvP01 genome. Computational studies have shown that the VIR genes from different *P. vivax* isolates can be grouped into a number of clusters, and it is possible that genes within clusters may be performing a similar function (30, 32). Cluster analysis showed that the majority of the 1145 PvW1 VIR proteins could be clustered into groups with VIRs from the PvP01, PvT01, PvC01 and SalI strains (**Fig. 6**), with no evidence that specific clusters are restricted to specific genomes or geographical regions.

**Figure 6.**
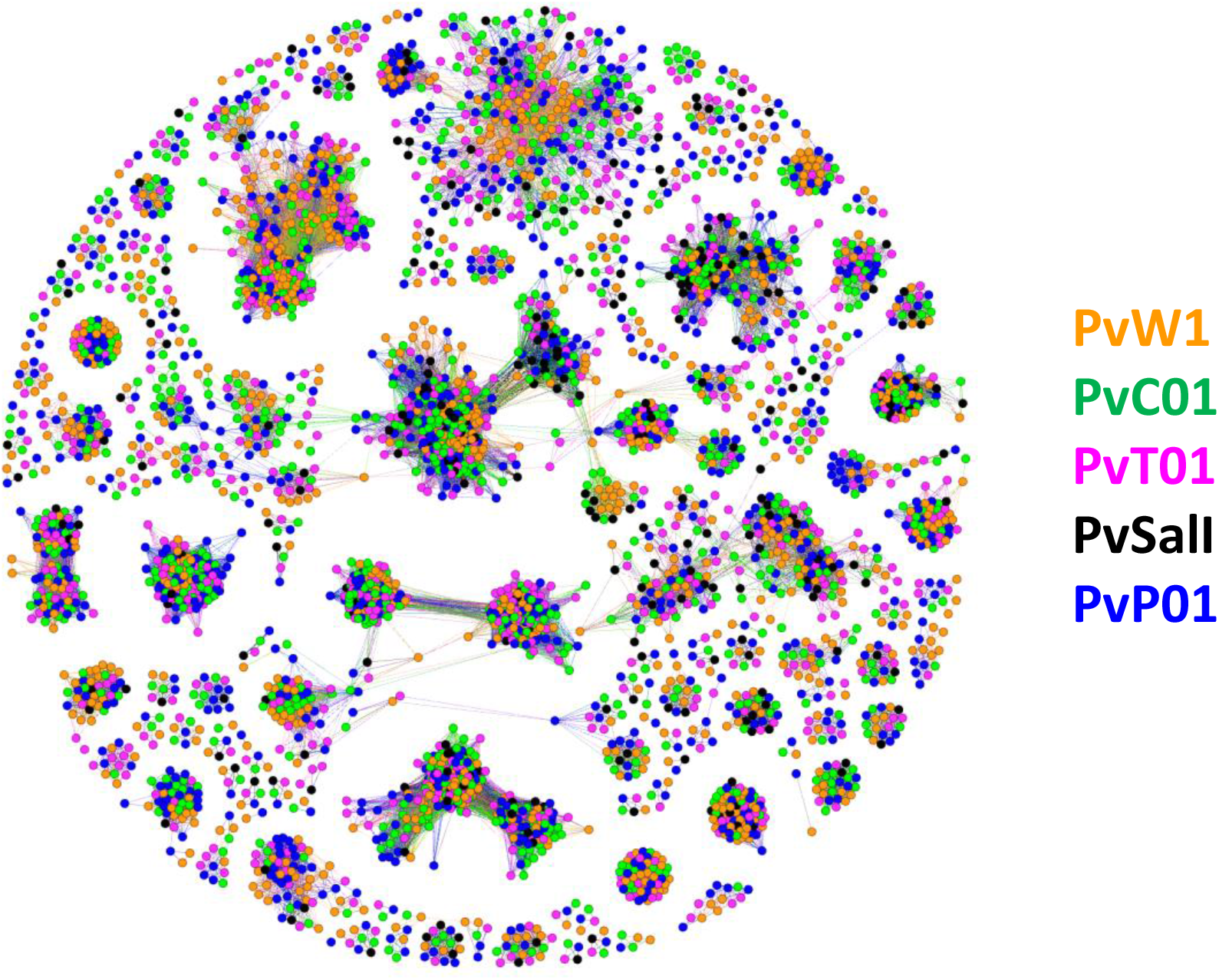
Cluster analysis of the PvW1 vivax interspersed repeat (VIR) proteins. Cluster analysis of the 1145 predicted VIR proteins encoded by the PvW1 genome compared to those of other *P. vivax* isolates (30, 31). Each spot represents a VIR protein from either PvW1 (orange), PvC01 (green), PvT01 (pink), PvSalI (black) and PvP01 (blue). Relatedness between the proteins is represented by distance, therefore more closely related proteins cluster together. Most of the clusters contain proteins from several isolates suggesting that the clusters are not restricted to specific genomes or geographical distribution.

Similarly, we resolved other smaller but still highly polymorphic multigene families such as the merozoite surface protein 3 (MSP3) family. These proteins are expressed on the surface of the invasive merozoite and are known to be highly polymorphic both in sequence and gene number between isolates. We compared the organization of the MSP3 multigene family in PvW1 to *P. vivax* isolates: PvP01 (30) and SalI, India-7, North Korean, Mauritania-1 and Brazil-1 (33). Genes flanking the MSP3 cluster (PVX_097665 and PVX_097740) are syntenic across all isolates, as are MSP3.1, MSP3.2, MSP3.3, MSP3.G, MSP3.10 and MSP3.11. There is however clearly variability in the central region of the MSP3 region, with MSP3.4, MSP3.5, MSP3.6, MSP3.7, MSP3.8 and MSP3.9 all present in some isolates but not others (**Fig. S7**). The arrangement of the PvW1 MSP3 cluster appears identical to that of PvP01.

### PvW1 vaccine candidate and drug resistance associated genes

The quality of the PvW1 genome also makes it easy to obtain and analyze potential vaccine targets, which we did for three high profile candidates (34), comparing the PvW1 sequence with those from PvP01 and SalI. The sporozoite-stage target circumsporozoite protein (PvCSP) is known to contain one of two major types of repeat called VK210 and VK247 (35, 36), with this heterogeneity an important factor for vaccine design. PvW1 contains VK210 repeats, the most prevalent form worldwide (**Fig. S8A**). The sequence of the transmission-stage candidate Pvs25 is highly conserved between PvW1 and other genomes, apart from the commonly variable amino acids 130 and 131 within the third epidermal growth factor (EGF)-like domain (**Fig. S8B**). Finally, we reviewed the PvDBP sequence given two vaccine candidates targeting region II are currently in early-phase clinical trials (29, 37). PvDBP in PvW1 has multiple polymorphisms with 10 in region II, including the DEK epitope (38), as compared to the SalI sequence used in the current clinical vaccines (29, 37). Like PvDBP from SalI, this gene in PvW1 also has a nine amino acid deletion (downstream of region II) that is not present in PvP01 (**Fig. S8C**). As well varying at a sequence level, PvDBP is also known to vary between isolates in copy number, with some isolates containing multiple copies (39, 40) now linked to evasion of humoral immunity (41). We therefore used Illumina read mapping across the PvW1 genome assembly to check for copy number variation of genes. Here, if regions of the genome are present in multiple copies then the read coverage over that region would be higher than the surrounding regions. There was no evidence for increased coverage at either PvDBP or its homologue PvDBP2 (also called *P. vivax* erythrocyte-binding protein, PvEBP), suggesting both are present at a single copy within the PvW1 genome (**Fig. S9A-B**). We also looked at an uncharacterized gene on chromosome 14, homologous to PVX_101445 / PvP01_1468200, which has been shown to be duplicated in some isolates (42). This gene is also present in a single copy in PvW1 (**Fig. S9D**).

Drug resistance is not as well characterized in *P. vivax* as in *P. falciparum,* but several genes and polymorphisms have been associated with resistance in field studies. We therefore examined the sequences of four genes within the PvW1 genome that have been associated with drug resistance: dihydrofolate reductase (*PvDHFR*), dihydropteroate synthetase (*PvDHPS*), chloroquine resistance transporter (*PvCRT*) and multidrug resistance transporter 1 (*PvMDR1*). The PvW1 *PvDHFR* gene encodes a protein with the quadruple mutation F57L/S58R/T61M/S117T that has been linked to pyrimethamine resistance (43), whereas *PvDHPS* showed no mutations previously associated with sulfadoxine resistance (44). The molecular basis of *P. vivax* chloroquine resistance is less clear, although there is some evidence that mutations in *PvCRT* (K10 insertion) and *PvMDR1* (Y976F mutation) may be involved (45–47). Neither of these mutations are present in the PvW1 *PvCRT* and *PvMDR1* genes. It is important to note that both Riamet® and Malarone antimalarials were used with 100% treatment success rates in the VAC068 and VAC069A studies (both volunteers in VAC068 and 5/6 volunteers in VAC069A received Riamet®, 1/6 received Malarone), and none of the polymorphisms identified have been associated with resistance to either of these drugs.

## Discussion

Here we undertook CHMI model development for *P. vivax* and established a new PvW1 clonal isolate from Thailand. Our methodology elected to focus on a mosquito-bite CHMI protocol to provide the initial source of blood-stage parasites for the cryopreserved stabilate. The main advantages here (over parasites donated by returning travellers) included the ability to control the parasite source, the recruitment of suitable healthy volunteers (especially with regard to health screening and universal donor blood group) and logistical timing. We also created the blood stabilate as close as possible to the mosquito-stage, with only ∼3 cycles of replication from the liver (given it is known that mosquitoes reset parasite virulence (24)). If parasites had been cryopreserved from returning travellers or chronically-infected adults in an endemic setting they would have been selected over many rounds of asexual replication *in vivo* before creating the stabilate. This diminishes the criticism that blood-stage CHMI is not the natural route of infection. Notably our real-time assessment of parasite genotypes in the infected mosquitoes in Thailand identified only one clonal infection out of three tested. In future it will likely be necessary to screen more infected patient samples if parasite clones with specific genotypes are desired. It is also probable that this clonal infection resulted from a single relapsing hypnozoite in the patient, given natural infections are frequently polyclonal, arising from primary infections with multiple genotypes and meiotic siblings produced in the mosquito and/or multiple heterologous hypnozoites relapsing at a similar time (48–50).

The VAC068 mosquito-bite trial demonstrated feasibility and safety of this CHMI model for the first time at a European site, albeit in only two healthy adult UK volunteers. Both were successfully infected, with parasites first detectable by qPCR on dC+8.5 and the first wave of blood-stage parasitemia peaking around dC+9. This is largely consistent with data from humanized mouse models suggesting that the complete maturation of *P. vivax* liver stages and exo-erythrocytic merozoite release occurs between days 9 and day 10 post-sporozoite infection (51). Growth of blood-stage parasitemia was subsequently similar in the two volunteers, with both meeting criteria to donate blood on dC+14, prior to radical cure treatment with Riamet® followed by primaquine. Both volunteers were screened to have CYP2D6 genotypes predicted to be extensive metabolizer phenotypes of primaquine, and as of ∼3 years’ long-term follow-up, no relapse of infection has been documented.

Cryopreservation of the iRBC stabilate was performed successfully, however, given *P. vivax* cannot be cultured long-term *in vitro* it proved challenging to confirm parasite viability following thaw of the frozen stabilate, especially given the relatively low level of parasitemia achieved by CHMI in non-immune adults. However, given the stabilate from Donor 1 showed demonstrable growth *in vitro* using a short-term culture assay, we elected to proceed with this material for onward testing. Poor parasite recovery from Donor 2 could be associated with the predominant lifecycle stage at the time of cryopreservation; here microscopy records indicated the presence of more schizonts and a smaller proportion of early ring-stage trophozoites in comparison to Donor 1. Previous evidence suggests that the late asexual intra-erythrocytic parasites are not viable after cryopreservation with glycerolyte (52), and this may have led to the poor recovery of live parasites in Donor 2’s stabilate.

Previous reports of blood-stage CHMI using *P. vivax* have used one vial of cryopreserved stabilate to infect one volunteer (19–21), in contrast to similar studies with the stabilate of 3D7 clone *P. falciparum* whereby a single vial is diluted and routinely used to infect ∼20-30 volunteers (16, 53). Thawing many vials to undertake CHMI in larger cohorts of volunteers, e.g. for vaccine efficacy trials, brings many practical difficulties and, in turn, more rapidly depletes the bank of cryopreserved stabilate which is a finite resource. Conserving vials and building up a long-term safety database of the challenge agent for future use across many clinical studies is also preferable. Consequently, we assessed three different doses of the PvW1 blood-stage inoculum in the VAC069A study, with two volunteers receiving each dose. All six volunteers were successfully diagnosed at similar levels of blood-stage parasitemia within 12-16 days. Importantly, these data suggest that blood-stage CHMI trials in larger volunteer cohorts are now practical and feasible, whilst preserving the bank of PvW1 parasites for the long-term.

The AE profiles of both the mosquito-bite and blood-stage CHMI with PvW1 were highly comparable to previous reports of both models in malaria-naïve/non-immune adults using other isolates of *P. vivax* at the Colombian (8–10) or Australian sites (19–23). No SAEs occurred in either trial and all drug treatments were successful. Symptoms consistent with malaria were experienced and peaked post-treatment prior to resolving within a few days. We also observed transient thrombocytopenia and lymphocytopenia, as well as rises in ALT six days post-treatment, consistent with the reports of other sites undertaking *P. vivax* CHMI (9, 10, 54) and with no apparent impact on volunteer safety. We also observed consistent sero-conversion to PvMSP1_19_ post-CHMI in all volunteers, as reported in the Colombian CHMI trials (10, 55), but no detectable responses to PvDBP_RII. These data are in line with our similar studies of *P. falciparum* CHMI, with sero-conversion of malaria-naïve adults observed to immuno-dominant merozoite surface proteins following primary acute malaria exposure, but not to more transiently exposed RBC invasion ligands (56, 57).

Following mosquito-bite CHMI we observed ∼10-fold growth in blood-stage parasitemia per 48 hours, consistent with other reports for *P. vivax* (20), as well as our experience with *P. falciparum* (16, 53). Interestingly, however, the average PMR was lower (∼5.5-fold growth per 48 hours) following blood-stage CHMI with the same parasite. There was no obvious effect of challenge dose or Duffy blood group sero-phenotype on the PMR, the latter consistent with our observations *in vitro* using *P. knowlesi* parasites transgenic for PvDBP (58). However, Duffy blood group sero-phenotype has been linked to susceptibility of *P. vivax* clinical malaria following natural infection (59). Consequently CHMI studies in larger numbers of volunteers will be required to more stringently assess for any relationships between blood group antigens and the observed PMR, and to more accurately establish the natural variability in the PMR observed in malaria-naïve adults. A second striking difference between the two CHMI models was the apparent minimal gametocytemia following mosquito-bite CHMI, in contrast to blood-stage CHMI. In the latter, the *pvs25* transcripts (a marker of mature female gametocytes) were reliably detected in all six volunteers, reaching comparable levels to those reported in other *P. vivax* blood-stage CHMI studies (19, 20). Notably, poor transmission to mosquitoes was reported in another *P. vivax* mosquito-bite CHMI trial, consistent with our data here (60). Interestingly, a more recent study comparing the same two CHMI models with *P. falciparum* reported the same finding (17). Why blood-stage CHMI appears to lead to much greater gametocytemia than mosquito-bite CHMI, despite reaching comparable levels of overall parasitemia by the time of diagnosis, remains to be determined. However, this might reflect the greater number of asexual growth cycles since liver egress, or a longer time to diagnosis allowing for an extended window for conversion of asexual parasites.

Finally, we proceeded to undertake a genomic analysis of the new *P. vivax* PvW1 clone. The need to drug treat volunteer infections at relatively low parasitemia limited the amount of PvW1 parasite DNA that could be isolated for sequencing. Nevertheless, a very high quality genome assembly for PvW1 was created by using a hybrid assembly method which combined long PacBio reads with short Illumina reads. The PacBio library was created using low-input PacBio technology developed to create a genome assembly from a single mosquito (61), and is to our knowledge the first time that this has been applied to *Plasmodium* parasites. Our goal is that the PvW1 clone will become a valuable tool for vaccine discovery, drug testing and assessment of *P. vivax in vivo* immuno-biology. Accurate assessment of both the sequence and copy number of vaccine candidate antigens within the PvW1 genome will thus be critical in designing future vaccine immunogens and interpreting CHMI efficacy studies. The high quality of the PvW1 assembly allowed us to easily report on leading vaccine candidate antigens, analyze genes and polymorphisms associated with drug resistance in field studies, and resolve 1145 VIR genes as well as the smaller polymorphic PvMSP3 multigene family. Although the function of the highly variable subtelomeric multigene VIR family is not well defined, related genes are found in high numbers in most *Plasmodium* species which infect humans, monkeys and rodents, and some are thought to be involved in immune evasion, including by directly binding to and down-regulating natural killer (NK) cell ligands (62). Our cluster analysis will now enable comparison of gene function within and between clusters, and should help in the future elucidation of the function of the VIR gene family.

In conclusion, we have developed a mosquito-transmitted stabilate using a new clonal field isolate of *P. vivax* and combined new methodologies for parasite isolation and ultra-low input PacBio sequencing to assemble a reference-quality genome for CHMI. This has i) revealed polymorphisms in leading drug and vaccine targets that can now be functionally tested *in vivo* with PvW1 and ii) used a hybrid PacBio/Illumina genome assembly technique to identify 1145 unique VIR genes. This will allow for *in vivo* switching and selection of multi-gene families to be measured in *P. vivax* in the same way as has been done for *P. falciparum* (18). This has allowed us to open up many new research avenues, and in the first instance, we have used this model to investigate myeloid cell activation, systemic inflammation, and the fate and function of human T cells during a first-in-life *P. vivax* infection (63). The PvW1 parasite should prove to be an invaluable resource for the wider malaria community.

## Data Availability

Requests for materials should be addressed to the corresponding authors.
PvW1 genomic assembly and annotation data can be found at:
ftp://ngs.sanger.ac.uk/scratch/project/pathogens/ea10/pvivax/renamed_scaffolds/

ftp://ngs.sanger.ac.uk/scratch/project/pathogens/ea10/pvivax/renamed_scaffolds/

## Acknowledgments

This work was funded in part by the European Union’s Horizon 2020 research and innovation programme under grant agreement 733073 for MultiViVax; the UK Medical Research Council (MRC) Confidence in Concept Scheme at the University of Oxford [MC_PC_16056]; and by the National Institute for Health Research (NIHR) Oxford Biomedical Research Centre (BRC). The views expressed are those of the authors and not necessarily those of the NIHR or the Department of Health and Social Care. CMN is a Wellcome Trust Sir Henry Wellcome Postdoctoral Fellow [209200/Z/17/Z]. PJS is the recipient of a Sir Henry Dale Fellowship jointly funded by the Wellcome Trust and the Royal Society [107668/Z/15/Z]. TAR held a Wellcome Trust Research Training Fellowship [108734/Z/15/Z]. FB is the recipient of a Wellcome Trust PhD studentship [203764/Z/16/Z]. AK, EA, TDO, AJR and JCR were supported by the Wellcome Trust [206194/Z/17/Z]. AMB is supported by the MRC [MR/N00227X/1], Isaac Newton Trust, Alborada Fund, Wellcome Trust ISSF and University of Cambridge JRG Scheme, GHIT, Rosetrees Trust and the Royal Society. SB and SJD are Jenner Investigators and SJD held a Wellcome Trust Senior Fellowship [106917/Z/15/Z].

The authors are grateful for the assistance of: Julie Furze, Duncan Bellamy, Richard Morter, Catherine Mair, Lola Matthews, Natalie Lella, Daniel Marshall-Searson, Kathryn Jones and Chris Williams (Jenner Institute Laboratories and CCVTM, University of Oxford); Richard Tarrant, Eleanor Berrie and Emma Bolam (Clinical Biomanufacturing Facility, University of Oxford); Julie Staves and the Hematology Department (Oxford University Hospitals NHS Foundation Trust); Anjali Yadava (WRAIR, USA), Richard Tedder (Imperial College London, UK) and Nick Day (MORU, Thailand) for clinical advice; Jake Baum (Imperial College London, UK) for support with mosquito-bite CHMI; Sally Pelling-Deeves and Carly Banner for arranging contracts (University of Oxford); Karl Hoyle for providing training (Applied Science, UK); Colin Sutherland (LSHTM, UK), Carole Long (NIAID, NIH, USA) and Chetan Chitnis (Pasteur Institute, France) for providing reagents; members of the Wellcome Sanger Institute DNA Pipelines team, particularly Mandy Sanders, Craig Corton and Karen Oliver for their advice and input into the DNA sequencing process; Chris Jacob, Sonia Goncalves and the MalariaGEN team for support with parasite genotyping; Wai-Hong Tham, Meta Roestenberg and Susan Barnett for providing scientific advice as part of the MultiViVax Scientific Advisory Board; and all the study volunteers.

## Author Contributions

- Conceived and performed the experiments: AMM, YT, SES, JRB, AK, DQ, CMN, NJE, TAR, FRL, WR, KJE, J-SC, TDO, AJR, FB, GML, IDP, AM, MZ, MM, RLR, MB, CHM, JCS, NR, CK, NM, CS, TP, DJR, AMB, PJS, JS, SB, JCR, SJD.
- Analyzed the data: AMM, YT, SES, JRB, AK, NJE, KJE, J-SC, EA, TDO, AJR, JCS, MMH, BK, JS, SB, JCR, SJD.
- Project Management: AML, FLN, KJJ, IJT.
- Wrote the paper: AMM, AK, SES, JRB, JCR, SJD.

## Data and Materials Availability

Requests for materials should be addressed to the corresponding authors.

PvW1 genomic assembly and annotation data can be found at: ftp://ngs.sanger.ac.uk/scratch/project/pathogens/ea10/pvivax/renamed_scaffolds/

## Methods

### Thailand: Source patient case-finding and preparation of infected mosquitoes

For infection of mosquitoes, source patients were recruited from a medical clinic in Songkhala, one of Thailand’s southern *Plasmodium vivax* endemic areas; this protocol was approved by the Ethical Committee of The Faculty of Tropical Medicine, Mahidol University, Thailand (protocol number TMEC 18-014). Patients were consented to having a 20 mL blood sample taken for blood-borne infection testing, endemic mosquito-borne infection testing and *P. vivax* diagnostic testing. *P. vivax* was first diagnosed by microscopy at the field site, then confirmed by microscopy and nested PCR analysis following transport of the blood samples to the Mahidol Vivax Research Unit (MVRU), Mahidol University, Bangkok, to rule out the presence of any other *Plasmodium* species (data not shown). As *Anopheles* species are also known vectors of *Wucherichia bancrofti*, the main causative agent of lymphatic filariasis, source patient blood was also screened for filarial disease via rapid diagnostic test for IgG4 antibodies to the *W. bancrofti* Wb123 antigen (Standard Diagnostics, Inc.); all tests were negative.

Approximately 5 mL of each 20 mL blood sample was used to feed up to 3000 laboratory-bred *Anopheles dirus* mosquitoes via a direct membrane feeding system at MVRU. These mosquitoes were previously reared in the laboratory and fed only on rigorously screened human blood (purchased from the Red Cross) to maintain the colony and induce egg-laying. Mosquito infectivity was confirmed at 6-7 days post-feeding via oocyst counts following dissection of the midgut from representative mosquitoes.

In parallel to the mosquito-feeding, and in real-time, the remaining ∼15 mL of the source patients’ serum and whole blood samples were shipped from Thailand to the UK. These underwent additional testing for blood-borne infections and mosquito-borne diseases other than malaria. For maximal assurance of safety, serological tests for human immunodeficiency virus-1 (HIV-1) and HIV-2, human T cell lymphotropic virus-1 (HTLV-1) and HTLV-2, hepatitis B and C and syphilis were performed at Oxford University Hospitals NHS Trust, Oxford, UK. Alongside these, whole blood from the source patients were also screened for Japanese B encephalitis virus and chikungunya virus by PCR at the Rare Imported Pathogens Laboratory (RIPL) in the UK, due to anecdotal reports of both arboviral infections in *Anopheles* species (64) and in line with the protocols followed by WRAIR in their previous *P. vivax* CHMI study (12). Finally, as a further precautionary measure, although *Anopheles* species are not known to be vectors of dengue, Zika or West Nile viruses, PCR for these infections were also performed at RIPL on the source patients’ blood samples. All source patient infection screen tests were negative.

Alongside infection testing, molecular speciation of *P. vivax* was re-confirmed for each sample by nested PCR using whole blood and an in-house research-grade laboratory assay method, adapted from (65), at the University of Oxford. Positive control DNA samples for different *Plasmodium* parasite species were a kind gift from Prof Colin Sutherland (LSHTM, UK). Each sample was also genotyped to measure multiplicity of *P. vivax* infection; here extracted DNA was processed by the Wellcome Sanger Institute in Cambridge, UK using a SNP barcode panel as described by the MalariaGEN network (66), with detailed methods available at https://www.malariagen.net/resource/29.

The batch of mosquitoes fed off blood from patient C05-001 was ultimately selected for use in the VAC068 CHMI trial, and was shipped from MVRU in Thailand to Imperial College London, UK. Shipment took less than 48 hours, and no mosquito mortality was observed.

### VAC068: Study approvals

VAC068 was a clinical study to assess the safety of controlled human *P. vivax* malaria infection through experimental sporozoite inoculation (by mosquito-bite) of healthy malaria-naïve UK adults, and to characterize parasite growth and immune responses. The study was conducted in the UK at the Centre for Clinical Vaccinology and Tropical Medicine (CCVTM), University of Oxford (follow-up post-CHMI, admission for blood donation and treatment) and at the Sir Alexander Fleming Building (Infection and Immunity section) Imperial College of Science, Technology and Medicine, London (sporozoite challenge of volunteers). Recruited volunteers were healthy, malaria-naïve adults (male and female) aged between 18 and 50 years. The trial was registered on ClinicalTrials.gov (NCT03377296) and was conducted according to the principles of the current revision of the Declaration of Helsinki 2008 and in full conformity with the ICH guidelines for Good Clinical Practice (GCP). All volunteers signed written consent forms, and consent was checked to ensure volunteers were willing to proceed prior to CHMI. The study received ethical approval from the UK NHS Research Ethics Service (Oxfordshire Research Ethics Committee A, Ref 17/SC/0389).

GCP compliance was independently monitored by the University of Oxford Clinical Trials and Research Governance (CTRG) Office. An independent local safety monitor and safety monitoring committee acted as independent experts, who, if required, could evaluate any adverse events and advise the Investigators on treating or referring a volunteer to secondary care.

The primary objectives of the trial were i) to assess the safety and feasibility of *P. vivax* sporozoite CHMI (via mosquito-bite) in two healthy human volunteers; ii) to assess the immune response to primary *P. vivax* infection delivered by mosquito bite; and iii) to asses gametocytemia following primary *P. vivax* infection delivered by mosquito bite. Secondary objectives were to obtain up to 250 mL of blood from each infected volunteer and produce a cryopreserved stabilate of parasite-infected human red blood cells (iRBC) for future use in blood-stage *P. vivax* CHMI studies.

### VAC068: Specific considerations for screening of healthy UK adult volunteers

Two healthy UK adult volunteers were consented and enrolled into the VAC068 trial. Alongside the routine screening and inclusion/exclusion criteria used for CHMI trials at the University of Oxford site (see below), these volunteers were also specifically screened to be: i) blood group O rhesus-negative (O-), i.e. universal donors suitable for production of the cryopreserved iRBC stabilate; ii) Duffy-blood group positive, to ensure successful *P. vivax* blood-stage infection (7, 25); iii) glucose-6-phosphate dehydrogenase (G6PD) normal, to ensure no hemolytic anemia following curative treatment with primaquine (26); iv) cytochrome P450 2D6 (CYP2D6) genotype predicted to be an extensive metabolizer phenotype (27), to minimize chance of primaquine drug treatment failure against hypnozoites (13), and v) able to satisfactorily metabolize primaquine after administration of a 30 mg test dose (13).

Blood group (ABO, Rhesus and Duffy) were characterized and G6PD activity levels were measured in the NHS Hematology Laboratory at Oxford University Hospitals NHS Trust, UK. CYP2D6 genotype testing was performed by PharmGenomics GmbH, Germany, using the GenoChip method and NCBI reference sequence NG_008376.3. Classification was done according to (27). Measurement of the pharmacokinetic parameters of primaquine was performed by the Division of Experimental Therapeutics, Drug Metabolism and Disposition, at the Walter Reed Army Institute of Research, USA. Plasma samples from both volunteers, taken from 0 to 24 h after a single dose of primaquine (PQ), were analyzed by Liquid Chromatography-Mass Spectrometry (LC-MS) for levels of PQ and its major metabolite, carboxyprimaquine (cPQ). All plasma samples were frozen at −80 °C in laboratory facilities until ready for analysis. Calibration and quality control samples were prepared by spiking blank human plasma with the analyte of interest. Calibration, quality control, and study samples were extracted using a 2x volume of acetonitrile containing an internal standard (mefloquine, MQ). The ratio of the peak area of the analyte to the peak area of the internal standard was used for calibration and interpolation of sample concentrations. LC-MS methodology has been previously described in detail (13, 67).

Each volunteer also underwent an extensive screen for blood-borne infections, performed in line with the Joint UK Blood Transfusion and Tissue Transplantation Services Professional Advisory Committee guidelines (68), in the microbiology laboratory at OUH NHS Trust. Testing comprised serological tests for HIV-1 and HIV-2, hepatitis B and C, syphilis (anti-treponemal antibody), and HTLV-1 and HTLV-2 at screening; and nucleic acid amplification tests for HIV-1 and hepatitis B and C, as well as repeat serological tests for HTLV-1 and HTLV-2 and syphilis 7 days before challenge. In addition, the volunteers were screened serologically for Epstein-Barr virus (EBV) and cytomegalovirus (CMV). However, given i) both of these viruses are cell-associated, being carried within leukocytes, and the risk of transfusion-induced CMV and EBV infection has been deemed to be minimal following leukodepletion (69); and ii) the historical experience of us and others with a *P. falciparum* 3D7 clone blood-stage inoculum that originated from a CMV and EBV seropositive donor (15, 16), we did not exclude volunteers based on their serostatus for these two viruses.

### VAC068: Other routine screening, and full list of inclusion and exclusion criteria

For the two VAC068 volunteers, a medical history and physical examination were conducted at the screening visit. Hematology screening bloods included a full blood count, and a hemoglobinopathy screen was performed retrospectively post-challenge; whilst biochemistry measurements at screening included urea and electrolytes, liver function tests, magnesium & cholesterol. Dipstick urinalysis for all volunteers and pregnancy testing for female volunteers were conducted at screening, as well as an electrocardiogram. Pregnancy testing (in the form of serum beta human chorionic gonadotrophin, BHCG) was also carried out in female volunteers the day before CHMI (dC-1), and then at 7 days post-CHMI (dC+7), dC+14, just prior to starting primaquine treatment and at dC+21. The full list of inclusion and exclusion criteria is shown below:

#### Inclusion criteria

Volunteers had to satisfy all the following criteria to be eligible for the study:

- Healthy adult aged 18 to 50 years.
- Blood group O, Rhesus negative.
- Red blood cells positive for the Duffy antigen/chemokine receptor (DARC).
- High metabolizer of primaquine (as determined by CYP2D6 genotype).
- Normal serum levels of glucose-6-phosphate dehydrogenase (G6PD).
- Satisfactory serum levels of primaquine (when administered as test dose).
- Able and willing (in the Investigator’s opinion) to comply with all study requirements.
- Willing to allow the Investigators to discuss the volunteer’s medical history with their General Practitioner.
- Women only: Must practice continuous effective contraception for the duration of the clinic visits (first 3 months post-CHMI).
- Agreement to refrain from blood donation during the course of the study and for at least 5 years after the end of their involvement in the study.
- Written informed consent to participate in the trial.
- Reachable (24/7) by mobile phone during the period between CHMI and completion of all antimalarial treatment.
- Willing to take a curative anti-malaria regimen following CHMI.
- Willing to be admitted to the research bay at the CCVTM on the Churchill Hospital, Oxford site for blood donation and clinical monitoring, until antimalarial treatment is underway and their symptoms are settling.
- Willing to reside in Oxford for the duration of the study, until all antimalarials have been completed.
- Answer all questions on the informed consent quiz correctly.

### Exclusion Criteria

Volunteers were not eligible to participate if any of the following applied:

- History of clinical malaria (any species).
- Travel to a clearly malaria endemic locality during the study period or within the preceding six months.
- Use of systemic antibiotics with known antimalarial activity within 30 days of CHMI (e.g. trimethoprim-sulfamethoxazole, doxycycline, tetracycline, clindamycin, erythromycin, fluoroquinolones and azithromycin).
- Blood group A/B and/or Rhesus positive.
- Red blood cells negative for the Duffy antigen/chemokine receptor (DARC).
- Glucose-6-phosphate dehydrogenase (G6PD) deficient.
- Inadequate serum levels of primaquine (when administered as test dose).
- Current anemia (hemoglobin < 9 g/dL).
- Use of immunoglobulins or blood products (e.g., blood transfusion) at any time in the past.
- History of sickle cell anemia, sickle cell trait, thalassemia or thalassemia trait or any hematological condition that could affect susceptibility to malaria infection.
- Venepuncture unlikely to allow a 250 mL blood donation (as determined by the Investigator).
- Receipt of an investigational product in the 30 days preceding enrolment, or planned receipt during the study period.
- Prior receipt of an investigational vaccine likely to impact on interpretation of the trial data or the *P. vivax* parasite as assessed by the Investigator.
- Any confirmed or suspected immunosuppressive or immunodeficient state, including HIV infection; asplenia; recurrent, severe infections and chronic (more than 14 days) immunosuppressant medication within the past 6 months (inhaled and topical steroids are allowed).
- History of allergic disease or reactions likely to be exacerbated by malaria infection.
- Pregnancy, lactation or intention to become pregnant during the study.
- Use of medications known to cause prolongation of the QT interval ***and*** existing contraindication to the use of Malarone.
- Use of medications known to have a potentially clinically significant interaction with Riamet® ***and*** Malarone.
- Any clinical condition known to prolong the QT interval.
- History of cardiac arrhythmia, including clinically relevant bradycardia.
- Disturbances of electrolyte balance, e.g. hypokalemia or hypomagnesemia.
- Family history of congenital QT prolongation or sudden death.
- Contraindications to the use of both of the proposed anti-malarial medications; Riamet® Malarone.
- Contraindications to the use of primaquine.
- History of cancer (except basal cell carcinoma of the skin and cervical carcinoma in situ).
- History of serious psychiatric condition that may affect participation in the study.
- Any other serious chronic illness requiring hospital specialist supervision.
- Suspected or known current alcohol abuse as defined by an alcohol intake of greater than 42 standard UK units every week.
- Suspected or known injecting drug abuse in the 5 years preceding enrolment.
- Hepatitis B surface antigen (HBsAg) detected in serum.
- Seropositive for HTLV-1 or -2 (antibodies to HTLV) at screening or at dC-7.
- Seropositive for hepatitis C virus (antibodies to HCV) at screening or at dC-7 (***unless*** has taken part in a prior hepatitis C vaccine study with confirmed negative HCV antibodies prior to participation in that study, and negative HCV RNA PCR at screening for this study).
- Seropositive for RPR (antibodies to syphilis) at screening or at dC-7.
- Detectable HIV or hepatitis C virus by PCR at dC-7.
- Positive family history in both 1^st^ AND 2^nd^ degree relatives < 50 years old for cardiac disease.
- Volunteers unable to be closely followed for social, geographic or psychological reasons.
- Any clinically significant abnormal finding on biochemistry or hematology blood tests, urinalysis or clinical examination. In the event of abnormal test results, confirmatory repeat tests will be requested.
- Any other significant disease, disorder, or finding which may significantly increase the risk to the volunteer because of participation in the study, affect the ability of the volunteer to participate in the study or impair interpretation of the study data.

### VAC068: Mosquito-bite CHMI at Imperial College, London

Sporozoite CHMI delivered by mosquito-bite was conducted in the designated category 3 (CL3) suite within the Sir Alexander Fleming Building (SAF) at Imperial College London, UK. Mosquitoes were supplied directly from Thailand via World Courier in a temperature-controlled box at 28 °C with receipt acknowledged by an appropriate researcher. Infected mosquitoes were immediately transferred to a secure room within the insectary and maintained on 10 % fructose until 24 hours prior to skin feeding. Prior to CHMI, an appropriately trained researcher prepared the mosquitoes into secure small pots that were brought to the volunteers as described in a Standard Operating Procedure (SOP). The two healthy UK adult volunteers screened and consented to take part in VAC068 were each exposed to five “infectious bites” under controlled conditions. Here, an infectious mosquito bite was defined post-skin feeding by microscopic examination, with confirmation of >10 sporozoites in the mosquito’s salivary glands as well as the presence of human blood in the midgut.

### VAC068: Participant follow-up post-CHMI

The two VAC068 volunteers were reviewed by telephone daily for the first 5 days post-CHMI (dC+1 to dC+5), then reviewed in clinic on the evening of day 6 post-CHMI (dC+6.5) and subsequently twice daily (morning/evening) to monitor for symptoms/signs of malaria and check for development of parasitemia by quantitative PCR (qPCR) and thick blood film microscopy.

Volunteers were admitted to the clinical trial unit at the CCVTM in Oxford according to a clinical / diagnostic algorithm. This stipulated admission at an absolute threshold parasitemia of >10,000 gc/mL OR at threshold of >2,000 gc/mL in the presence of significant malaria symptoms. Following admission, the protocol allowed for a 72 hour window in which to donate blood; volunteer 01-008 donated blood first at dC+14 and volunteer 01-004 at dC+14.5. A 250 mL blood sample was collected using aseptic technique, via a whole blood donation kit containing an in-line leukodepletion filter (Leuokotrap WB, Haemonetics Corp), at room temperature. The blood donation kit was placed on an agitator (Blood Collection Monitor and Mixer, HemoFlow 400^TM^, Applied Science UK Ltd). This maintained automatic mixing of the blood during flow at a rate of >30mL/min, ensuring combination with the contained anticoagulant and so minimizing the risk of coagulation. In order to anonymize the blood donor, these samples were randomized and relabelled either “Donor 1” or “Donor 2”. Traceability of the blood donor is, however, maintained within a confidential clinical record which may be accessed by the trial Chief Investigator on behalf of the trial Sponsor if deemed necessary for safety reasons.

Antimalarial treatment (60-hour course of artemether/lumefantrine, Riamet®) was started immediately after blood donation, followed by a 14-day course of primaquine, 30 mg once daily. Both volunteers attended clinic on alternate mornings for directly observed primaquine treatment (telephoned to confirm consumption on intervening days) until they had completed the course.

Final follow-ups in clinic were performed at dC+45 and at dC+90, and between these days volunteers were contacted fortnightly by email (on dC+59 and dC+73) to ensure they remained well and asymptomatic. Both volunteers also underwent repeat serological testing for HIV-1, HIV-2, hepatitis B and C, syphilis, HTLV-1 and HTLV-2 at dC+90 to ensure that no seroconversion from a recently-acquired infection (that may have been undetectable around the time of CHMI) had occurred since the challenge period. Subsequently the volunteers received an email from the Investigators fortnightly up until one year post-CHMI and then annually from 1-5 years post-CHMI (ongoing). This was to enquire about the presence of any symptoms suggestive of *P. vivax* malaria relapse (or any medical intervention sought/received) since they were last seen.

### Total parasite quantification

Quantitative PCR (qPCR) was used to monitor total *P. vivax* blood-stage parasitemia in volunteers’ blood in real-time. The assay targets the 18S ribosomal RNA (rRNA) gene and was adapted from previously published methodology (19, 53). DNA was initially extracted from 0.4 mL whole EDTA blood using the Qiagen DSP DNA Blood mini Kit. 5 % of each extraction (total eluate volume = 100 μL, with 5 μL used per assay) was run in triplicate for qPCR; equivalent to 60 μL blood directly assessed. An additional extraction was performed post-CHMI on aliquots of frozen blood from all time-points, using a QIAsymphony SP robot, utilizing the Qiagen DSP Blood Midi Kit and the pre-loaded Blood 400 v6 extraction protocol, with a 100 μL elution in ATE buffer selected, (giving identical extraction and elution volumes in both manual and automated extractions). Both methods had been shown to be equivalent, but with greater ease of use and reduced chance of cross contamination with the automated extraction. Additionally, aliquots of dC-1 samples were spiked with a known concentration of positive control DNA to check there was no presence of PCR inhibitors in volunteers’ blood prior to CHMI.

Following DNA extraction, a standard Taqman absolute quantitation was used against a standard curve to amplify a 183 bp PCR product from the multi-copy, highly conserved 18S ribosomal RNA genes of *Plasmodium* spp. qPCR using the following adapted oligonucleotide primers and probe (19): 18s forward primer 5’-AGG AAG TTT AAG GCA ACA ACA GGT-3’, 18s reverse primer 5’-GCA ATA ATC TAT CCC CAT

CAC GA-3’ and shortened FAM labelled probe sequence 5’-TGA ACT AGG CTG CAC GCG-3’, was run on an ABI StepOne Plus machine with v2.3 software. Default Universal qPCR (target FAM-NFQ-MGB) and QC settings were used apart from the use of 40 cycles and 25 μL reaction volume.

This qPCR detects DNA from pan-*Plasmodium* species, but unlike the synchronous growth of *P. falciparum*, circulating *P. vivax* iRBC may contain up to 10-15 individual genomes (in blood-stage late trophozoites and schizonts) and can also include the presence of gametocytes. The qPCR score is therefore reported in genome copies/mL (gc/mL) as opposed to a quantity of parasites.

The standard curve was generated from dilution of a linearized plasmid encoding part of the *Plasmodium* spp. 18S ribosomal RNA gene and calibrated using known *P. falciparum* spiked blood samples initially and then reference DNA extracted from whole blood from *P. vivax*-infected patient samples in Thailand where parasites had been quantified by microscopy (kindly provided by Mahidol University). Based upon earlier results obtained using dilution series of microscopically-counted cultured *falciparum* (Pf) parasites, a Pf-specific 18S rRNA Taqman qPCR showed a lower limit of quantification (LLQ, defined as %CV <20%) of around 20 Pf parasites (p)/mL blood (28). Counted parasite dilution series results also suggested that the lower limit of probable detection (LLD, i.e. a probability of >50% of ≥1 positive result among three replicate qPCR reactions) is in the region of 5 p/mL, while samples at 1 p/mL are consistently negative (24/24 qPCR reactions). Positive results in this assay (even at very low level) are thus essentially 100 % specific for genuine parasitemia, with positive results beneath the LLQ likely to signify parasitemia in the range 2-20 p/mL. Similar sensitivity in terms of genome copy detection was observed when using the pan-*Plasmodium* qPCR described above and the diluted *P. vivax*-infected patient blood test samples from Thailand. As noted, these samples had microscopically mixed life stages with varying copies of the 18S rRNA gene and thus the assay readout is reported in terms of gc/mL. Based on this and the above experiments, 20 gc/mL was set as the minimum level to meet positive reporting criteria, but all raw data are shown in the Results.

For quality control purposes, qPCR samples were re-tested if;

- Replicates included a mixture of positive and negative (in terms of amplification) results with one or more positive results > 100 gc/mL;
- The % CV of any results were high outliers.

All ‘passed’ data following the quality control steps above, including any 0 values, were used to generate the final mean qPCR result for each time-point.

### Thick blood film microscopy

Collection of blood, preparation of thick films and slide reading were performed according to Jenner Institute Standard Operating Procedure (SOP) ML009. Briefly, slides were prepared using Field’s stain A and then Field’s stain B. 200 fields at high power (1000x) were read. Visualization of 2 or more parasites in 200 high power fields constituted a positive result. For internal quality control, all slides were read separately by two experienced Thai microscopists, with a third read if results were discordant.

### VAC068: Cryopreservation and *in vitro* testing of *P. vivax* infected blood

After blood donation, the leukodepleted blood from both volunteers was maintained at ∼37 °C and transported immediately to the Jenner Institute Laboratories, University of Oxford. Here, RBC were separated from plasma by centrifugation before mixing the RBC with Glycerolyte 57 (Fenwal 4A7833) at 1:2 volume ratio. All procedures were conducted according to SOPs under stringent Quality Assurance (QA) oversight and guidance from a Qualified Person (QP) at the University of Oxford. The RBC-Glycerolyte mixture was finally aliquoted at 1.5 mL per cryovial, transferred into CoolCells (Corning 432009) and placed at −80 °C within 2 h 30 min of blood donation to freeze overnight; the following day the frozen cryovials were transferred to long-term storage in liquid nitrogen.

A final screen for blood-borne infections was conducted on the plasma, derived directly from the blood donation (separated from the RBC prior to cryopreservation), in line with testing procedures performed by the UK NHS Blood Transfusion service. RNA PCR for HIV-1 and hepatitis C, DNA PCR for hepatitis B, EBV CMV, and serology for HIV-2, HTLV-1, HTLV-2, and *Treponema pallidum* was performed on thawed plasma samples at University Hospitals Birmingham NHS Foundation Trust, UK (Public Health England, Birmingham Laboratory). Separately, screening of a blood sample from Donor 1 for the Kell blood group antigen was performed by Oxford University Hospitals NHS Trust Haematology Laboratory, UK.

The cryopreserved stabilate from Donor 1 was also tested for sterility by direct inoculation and mycoplasma by specific culture; both tests were negative. In addition, endotoxin was quantified by kinetic chromogenic limulus amoebocyte lysate assay, reporting a result <2 EU/mL. These assays were conducted by a Contract Research Organization: SGS Vitrology, Glasgow, UK or SGS Vitrology’s contracted services at Moredun Scientific, Penicuik, Scotland, UK. The tests were non-regulatory standard and performed for information only.

### Parasite viability assay

A vial of the cryopreserved stabilate was gently warmed at 37 °C, before addition of 0.2x volume 12 % NaCl dropwise. After 5 min incubation, the cells were pelleted at 1500 x*g* for 5 min. Supernatant was removed, before addition of 10x volume 1.6 % NaCl dropwise followed by 10x volume 0.9 % NaCl. The cells were then immediately spun down as before and the pelleted RBC transferred to short term *in vitro* culture. Here, the sample was added to McCoy 5A medium (Sigma) supplemented with 2.4 g/L D-glucose, 25 mM HEPES and 200 mM hypoxanthine (all from Sigma, St. Louis) and 20 % heat-inactivated human O serum, in an atmosphere of 5 % O_2_ at 37.5 °C. At each time point, a 20 µL aliquot of RBC was taken for DNA extraction and parasite genome copies quantified by qPCR. In addition, 10 µL RBC were used to make thick and thin blood smears for parasite growth and morphology monitoring by light microscopy. Here, Giemsa stain (Sigma, St. Louis) was filtered using a 0.8 µm filter (Merck Millipore, Ireland) then diluted to 5 % using water, prior to staining the slides at RT for 20 min before gently washing and drying the slides

### VAC069: Study approvals

The subsequent VAC069 trial assessed the safety and feasibility of blood-stage *P. vivax* CHMI through experimental inoculation with the cryopreserved PvW1 infected erythrocytes (collected in VAC068) in healthy malaria-naïve UK adults. The VAC069 trial is an on-going multi-part study, and the work reported here covers the first part of this trial in six volunteers (termed “VAC069A”). All six volunteers were challenged and followed up at the CCVTM, Oxford, UK. The trial was registered on ClinicalTrials.gov (NCT03797989) and was conducted according to the principles of the current revision of the Declaration of Helsinki 2008 and in full conformity with the ICH guidelines for GCP. All volunteers signed written consent forms, and consent was checked to ensure volunteers were willing to proceed prior to CHMI. The study received ethical approval from the UK NHS Research Ethics Service (South Central – Hampshire A Research Ethics Committee), Ref 18/SC/0577. Independent safety monitoring and GCP compliance was monitored as for VAC068.

The VAC069A trial tested safety and infectivity of the PvW1 cryopreserved stabilate (from Donor 1 in VAC068) by blood-stage CHMI, in line with prior experience using the *P. falciparum* blood-stage CHMI model (14–16). This proof-of-concept clinical trial sought to assess feasibility of infection at three different doses of PvW1 blood-stage inoculum. Two volunteers receive a whole vial’s worth of iRBC (“neat”), two volunteers received one fifth of the challenge dose via a 1:5 dilution, and the final two volunteers were inoculated with one twentieth of the dose via a 1:20 dilution.

### VAC069A: Study population and screening

This study recruited healthy, malaria-naïve adult volunteers (male and female) aged between 18 and 50 years. The inclusion and exclusion criteria for the VAC069A study were very similar to those of VAC068, with the only differences being removal of criteria related to ABO/Rhesus blood group, G6PD activity, CYP2D6 genotype and primaquine metabolism (as not relevant to blood-stage CHMI) and removal of viral serology at dC-7 (as these volunteers were not donating blood for use in future clinical studies). Duffy blood group positivity (and serological phenotype) was also confirmed. Screening hemoglobin cut-offs were also more conservative (Hemoglobin <120 g/L for a female volunteer or <130 g/L for a male volunteer prior to primary CHMI) to minimize the chances of anemia resulting from cumulative blood volume taken over the whole VAC069 trial period.

### VAC069A: Blood-stage inoculum preparation and CHMI

The PvW1 blood-stage inoculum was thawed and prepared under strict aseptic conditions as previously described for *P. falciparum* (16), with some modifications. Briefly, five vials of cryopreserved erythrocytes (containing approximately 0.5mL of red blood cells each) were thawed in parallel in a derogated containment level III laboratory area using solutions licensed for clinical use and single-use disposable consumables. A class II microbiological safety cabinet (MSC) was used to prepare the inoculum, which was fumigated with hydrogen peroxide and decontamination validated prior to use. To prepare the inoculum, 0.2 volume 12 % saline was added dropwise to the contents of each (∼1.5 mL) vial of thawed infected blood. Each sample was left for 5 min, before an additional 10 volumes of 1.6 % saline was added dropwise prior to centrifugation for 4 min at 800 x*g*. Each supernatant was removed, and 10 mL of 0.9% saline was added dropwise. The cell pellets were then washed twice in 0.9 % saline before a final resuspension in 0.9 % saline. At this final step, the five samples (from the five original cryovials) were combined into one 10 mL sample in 0.9 % saline. This 10 mL suspension was then divided into aliquots, equivalent to one original cryovial (i.e. 2mL), one fifth of a cryovial, and one twentieth of a cryovial by further dilution in 0.9 % saline. Each dosing aliquot was made up to a total volume of 5 mL in % saline in a sterile syringe for injection and transported to the clinic. Retrospective qPCR analysis indicated the neat inoculum dose to contain 2322 gc in total (although this is likely to contain a mixture of live and dead parasites post-thawing).

The reconstituted blood-stage inoculum (5 mL per syringe) was injected intravenously via an indwelling cannula, preceded and followed by a saline flush. Single volunteers from each of the three dosing groups were administered the inoculum first. The inoculum was subsequently administered to the remaining three volunteers (one from each dosing group). All six volunteers were inoculated within 2 h 24 min of thawing the cryopreserved stabilate. Volunteers were then observed for 1 h before discharge from the clinical facility. Following CHMI, a leftover sample of the inoculum was also cultured and shown to be negative for bacterial contamination.

### VAC069A: Participant follow-up post-CHMI

The VAC069A volunteers were reviewed in clinic once in the morning of day 1 post-CHMI (dC+1), then twice daily from day 2 until day 12 post-CHMI inclusive (dC+2 to dC+12.5). From dC+13 to dC+20.5, visits were either once or twice daily depending on the qPCR result. Once a qPCR threshold of 1000 gc/mL was reached, visits continued twice daily. If qPCR had not reached this threshold, visits reduced to once daily. Diagnostic criteria were based on thick blood film microscopy results and qPCR in the presence or absence of symptoms:

- If symptomatic: ≥2 parasites visible on 200 fields (thick blood film microscopy) OR a parasitemia of >5,000 gc/mL by qPCR.
- If asymptomatic: a parasitemia of >10,000 gc/mL on qPCR OR a parasitemia of >5,000 gc/mL PLUS ≥2 parasites visible on 200 fields (thick blood film microscopy).

Treatment was completed with either a 60-hour course of Riamet® or a 48-hour course of Malarone. Volunteers were directly observed taking their 24- and 48-hour doses (T+1 and T+2 visits, respectively). Subsequent follow-ups visits in clinic were 6 days post-initiation of antimalarial treatment (T+6), 28 days, 45 days and 90 days post-CHMI.

### VAC068 and VAC069A safety analysis

Data on both solicited AEs occurring during and after the CHMI period (that may have related to CHMI or antimalarial treatment) as well as any unsolicited AEs, were collected at clinic visits, from dC+1 up until the end of primaquine antimalarial treatment (VAC068) and until 6 days post-initiation of Riamet®/Malarone treatment (VAC069A). Volunteers were given a card on which to document the end date of any outstanding malaria symptoms on-going between completing anti-malarial therapy and their next clinic visit (dC+45 in VAC068, and dC+28 in VAC069A).

Data on serious adverse events (SAEs) were collected throughout the entire study period (5 years for VAC068, 3 months for VAC069A).

Volunteers graded all AEs as mild, moderate or severe:

- **GRADE 0:** None.
- **GRADE 1:** Transient or mild discomfort (< 48 h); no medical intervention/therapy required.
- **GRADE 2:** Mild to moderate limitation in activity – some assistance may be needed; no or minimal medical intervention/therapy required.
- **GRADE 3:** Marked limitation in activity, some assistance usually required; medical intervention/therapy required; hospitalization possible.

For each unsolicited AE, an assessment of the relationship of the AE to the study intervention (CHMI/ antimalarial treatment) was undertaken. Alternative causes of the AE, such as the natural history of pre-existing medical conditions, concomitant therapy, other risk factors and the temporal relationship of the event to the study intervention were considered. The likely causality of all unsolicited AEs was assessed as per the criteria below:

- **No Relationship:** No temporal relationship to study intervention ***and*** alternate aetiology (clinical state, environmental or other interventions); ***and*** does not follow known pattern of response to study intervention.
- **Unlikely:** Unlikely temporal relationship to study intervention ***and*** alternate aetiology likely (clinical state, environmental or other interventions) ***and*** does not follow known typical or plausible pattern of response to study intervention.
- **Possible:** Reasonable temporal relationship to study intervention; ***or*** event not readily produced by clinical state, environmental or other interventions; ***or*** similar pattern of response to that seen with other similar interventions.
- **Probable:** Reasonable temporal relationship to study intervention; ***and*** event not readily produced by clinical state, environment, or other interventions ***or*** known pattern of response seen with other similar interventions.
- **Definite:** Reasonable temporal relationship to study intervention; ***and*** event not readily produced by clinical state, environment, or other interventions; ***and*** known pattern of response seen with other similar interventions.

AE data also included the results of hematology (full blood count) and biochemistry (liver function tests, urea and electrolytes) carried out at dC+9, dC+11, within 12 hours of starting antimalarials after blood donation (C+14-14.5), and then at dC+45 and dC+90 (VAC068) and at dC+14, day of diagnosis, T+1, T+6, dC+28 and dC+90 (VAC069A).

### Gametocyte quantification

*P. vivax* gametocytemia was determined by one-step quantitative reverse transcription PCR (qRT-PCR) targeting the messenger RNA marker of female mature gametocytes, *pvs25*. For RNA extraction samples were processed within 4 h of blood sampling: here 50 μL whole blood was mixed with 250 μL RNA protect reagent (Qiagen) for RNA stabilization, until the blood was lysed and had turned black. Samples were then stored at −20 °C. Subsequently, 600 μL D-PBS with 1 % β-ME was added to each sample and then centrifuged for 15 min at 15,000 x*g*, the supernatant removed and the pellet resuspended in 300 μL RLT lysis buffer (Qiagen) + 1 % β-ME with 20 μL proteinase K and incubated at 55°C for 10 min. The samples were homogenized into a QIAshredder column (Qiagen) and purified through an RNeasy mini spin column (Qiagen) as per the manufacturer’s instructions. Each sample was then eluted in 100 μL RNAse-free water. Thereafter, one-step RT-PCR was performed using Luna® Universal Probe One-Step RT-qPCR Kit (New England Biolabs). Briefly, 5 μL RNA extract was added to a final reaction volume of 25 μL consisting of 12.5 μL Luna® Universal qPCR Master Mix, 0.625 μL Pv25 MGB-FAM probe (5’-CCA ATC CAG AAG ATG AGA-3’), 1.25 μL of each primer (diluted at 10 μM), and 3.375 μL nuclease-free water and 1 μL reverse transcriptase (RT) enzyme (NEB). The *pvs25* primer sequences were 5’-GTT GCT CAT GTG CTA TTG-3’ for the forward primer and 5’-CAG ACT TCA TTA TCT GTG TTA-3’ for the reverse primer. Analyses were performed on a StepOne Plus machine (Thermo Fisher Scientific) using the StepOne software v2.3. The thermal conditions consisted of reverse transcription (55 °C for 10 min), enzyme activation (95 °C for 1 min) and two-temperature cycling steps (95 °C for 10 s, 60 °C for 1 min, for 45 cycles). All samples were tested in triplicate. Controls without RT enzyme were added to exclude false positives due to the presence of genomic DNA and were tested in duplicate. Ct values were converted into *pvs25* transcript/µL using plate-specific standard curves, generated by serial-diluted *pvs25* RNA transcripts (10^7^ -10 copies/μL). Final values were multiplied by 2 (dilution factor at the RNA extraction step) to report per µL of the original blood sample. *pvs25* RNA transcripts were produced by amplification of the *pvs25* gene by T7 polymerase (HiScribe T7 High Yield RNA Synthesis Kit); RNA purified by lithium chloride extraction before treatment with DNAse I (Qiagen) to eliminate residual DNA; standards were diluted in DNA-free H_2_O containing a background of 5 % aspecific human RNA (isolated from blood of a healthy donor), to improve linearity of the standard curve.

### Modelling of PMR – VAC068

A qPCR-derived parasite multiplication rate (PMR) was modelled based on previously described methodology (28, 53, 70). In brief, to model the PMR, the arithmetic mean of the three replicate qPCR results obtained for each individual at each time-point was used for model-fitting. Negative individual replicates were assigned a value of 0 gc/mL for the purposes of calculating the arithmetic mean of triplicates (where at least one of the three readings was positive). All qPCR data points which, based upon the mean of the three replicates, were >5 gc/mL were used for modelling and any values ranging from 1-5 gc/mL were replaced with 5 gc/mL. The time interval between the morning and evening bleeds used for qPCR monitoring was set as 0.3 days. PMR per 48 hours was then calculated using a linear model fitted to log_10_-transformed qPCR data.

### Modelling of PMR – VAC069A

Analysis of VAC069A was performed as for VAC068 but additionally any data point that was negative but preceded a positive data point was replaced with a value = 5 gc/mL; otherwise negative data points occurring after any positive data point but not preceding a positive data point were treated as 0 gc/mL. The time interval between the morning and evening bleeds used for qPCR monitoring in this study was set as 0.37 days. PMR per 48 hours was then calculated using a linear model fitted to log_10_-transformed qPCR data.

### Peripheral blood mononuclear cell (PBMC), plasma and serum preparation

Blood samples were collected into lithium heparin-treated vacutainer blood collection systems (Becton Dickinson, UK). PBMC were frozen in foetal calf serum (FCS) containing 10% dimethyl sulfoxide (DMSO) and stored in liquid nitrogen. Plasma samples were stored at −80 °C. For serum preparation, untreated blood samples were stored at room temperature (RT) and then the clotted blood was centrifuged for 5 min (1000 *xg*). Serum was stored at −80 °C.

### Anti-PvDBP_RII standardized ELISA

ELISAs to quantify circulating PvDBP_RII-specific total IgG responses were performed using standardized methodology, similar to that previously described (29). Day C-1 and dC+90 serum or plasma samples from the VAC068 and VAC069A volunteers were tested, alongside samples from 8 healthy UK adults previously vaccinated in the VAC051 Phase Ia trial of a candidate PvDBP_RII vaccine (Group 2C) (29). Nunc MaxiSorp ELISA plates (Thermo Fisher) were coated overnight (≥16 h) at 4 °C with 50 µL per well of 2 µg/mL PvDBP_RII (SalI allele) protein (29). Plates were washed 6x with 0.05 % PBS/Tween (PBS/T) and tapped dry. Plates were blocked for 1 h with 100 µL per well of Starting Block™ T20 (Thermo Fisher) at 20 °C. Test samples were diluted in blocking buffer (minimum dilution of 1:100), and 50 µL per well was added to the plate in triplicate. Reference serum (made from a pool of high-titer vaccinated donor serum) was diluted in blocking buffer in a three-fold dilution series to form a ten-point standard curve. Three independent dilutions of the reference serum were made to serve as internal controls. The standard curve and internal controls were added to the plate at 50 µL per well in duplicate. Plates were incubated for 2 h at 20 °C and then washed 6x with PBS/T and tapped dry. Goat anti-human IgG–alkaline phosphatase secondary antibody (Merck) was diluted 1:1000 in blocking buffer and 50 µL per well was added. Plates were incubated for 1 h at 20 °C. Plates were washed 6x with PBS/T and tapped dry. 100 µL per well of PNPP alkaline phosphatase substrate (Thermo Fisher) was added, and plates were incubated for approximately 15 min at 20 °C. Optical density at 405 nm (OD_405_) was measured using an ELx808 absorbance reader (BioTek) until the internal control reached an OD_405_ of 1.0. The reciprocal of the internal control dilution giving an OD_405_ of 1.0 was used to assign an arbitrary unit (AU) value of the standard. Gen5 ELISA software v3.04 (BioTek) was used to convert the OD_405_ of test samples into AU by interpolating from the linear range of the standard curve fitted to a four-parameter logistic model. Any test samples with an OD_405_ below the linear range of the standard curve at the minimum dilution tested were assigned a minimum AU value of 5.0.

### Anti-PvMSP1_19_ ELISA

Anti-PvMSP1_19_-specific total IgG responses were measured in VAC068 and VAC069A volunteer serum and plasma via indirect ELISA (same test samples as for the PvDBP_RII ELISA). Nunc MaxiSorp ELISA plates (Thermo Fisher) were coated with 50 µL per well of 2 µg/mL PvMSP1_19_ protein (kindly provided by Dr Chetan Chitnis (71)) and left overnight (≥16 h) at 4 °C. Plates were washed 6x with 0.05% PBS/T and tapped dry. Plates were blocked for 1 h with 100 µL per well Starting Block™ T20 (Thermo Fisher) at 20 °C. Test samples were diluted 1:100 in blocking buffer and 50 µL per well was added in duplicate. A 1:6400 dilution of a post-CHMI positive control serum sample was also added in duplicate. Plates were incubated for 2 h at 20 °C and then washed 6x with PBS/T and tapped dry. Goat anti-human IgG–alkaline phosphatase secondary antibody (Merck) was diluted 1:1000 in blocking buffer and 50 µL per well was added. Plates were incubated for 1 h at 20 °C, then washed 6x with PBS/T and tapped dry. 100 µL per well of PNPP alkaline phosphatase substrate (Thermo Fisher) was added and plates were incubated for approximately 20 min at 20 °C. OD_405_ was measured using an ELx808 absorbance reader (BioTek) until the positive control reach an OD_405_ of 1.0. Results are plotted as the mean OD_405_ reading for each test sample.

### Illumina sequencing

Blood samples were used from the two volunteers in VAC068 as follows: first volunteer = 1 x 10 mL packed RBC (dC+11), plus 1 x 1.5 mL + 1 x 2 mL packed RBC (dC+14); second volunteer = 1 x 10 mL packed RBC (dC+11), plus 2 x 2.5 mL packed RBC (dC+14). DNA was extracted using the Qiagen blood DNA midi kit and sequenced with Illumina HiSeq X10 with 150 bp paired end reads.

### Long read sequencing

Preparation of schizonts: For the preparation of high molecular weight DNA for long-read sequencing blood samples were collected at diagnosis from volunteers enrolled in VAC069A and used to culture schizonts *ex vivo*. This approach was chosen to maximize the quantity of parasite DNA available. Briefly, 20 mL whole blood was depleted of leukocytes using NWF filters. RBC were then washed in McCoy’s medium and resuspended at 3 % hematocrit. Parasite growth medium (McCoy’s) was supplemented with 20 % AB human serum, 2.4 mg/mL D-glucose, 25 mM HEPES, 0.2 mM hypoxanthine and flasks were gassed with 5% O_2_, 5% CO_2_ (in N_2_) and incubated at 37 °C. The duration of culture was adapted to allow schizont maturation, based on the dominant life cycle stage present at the start of culture, as determined by Giemsa-stained thin and thick blood smears. Twelve hours prior to end of culture, the protease inhibitor E64 was added at a final concentration of 10 μM to prevent schizont rupture. At the end of the culture red cells were lysed and parasites isolated according to our previously published protocol: https://dx.doi.org/10.17504/protocols.io.brgjm3un. In brief, RBC were washed in PBS and lysed in 0.0075 % saponin for 10-15 min on ice; samples were then centrifuged at 2000 x*g* to pellet schizonts, which were snap-frozen on dry ice; and the supernatant was centrifuged at 18,000 x*g* to pellet less mature parasites (rings and trophozoites) – these were also snap frozen on dry ice. In the following steps for DNA extraction, samples containing different parasite stages were pooled to maximize the yield, and samples from all volunteers (except 01-003) were taken forward for sequencing.

High molecular weight DNA extraction: PvW1 parasite pellets were pooled into four groups. Each group was thawed on ice, resuspended in 200 µL cold PBS and extracted using the Qiagen MagAttract® HMW DNA Kit (blood protocol). This yielded a total of 107 ng high molecular weight DNA with an average fragment size of 78 kbp as measured by Femto Pulse system (Agilent).

Shearing and PacBio library construction and sequencing: The pooled high molecular weight DNA (107 ng) was sheared using a Diagenode Megaruptor 3 (speed setting 30) to an average fragment size of 18.2 kbp. SMRTbell® library preparation and clean up were as described in the manufacturer’s protocol for low input DNA: https://www.pacb.com/wp-content/uploads/Procedure-Checklist-Preparing-HiFi-Libraries-from-Low-DNA-Input-Using-SMRTbell-Express-Template-Prep-Kit-2.0.pdf. After a 1.8x Ampure PB bead clean-up to remove fragments below 250 bp, 87 ng PvW1 DNA remained for library preparation, which produced 30 ng of SMRTbell® library for sequencing. The SMRT libraries were sequenced on a single Sequel SMRT Cell 1M and yielded 13 Gb sequence (3.4 Gb unique).

### Raw sequencing data

Data are included in the study entitled “PvW1 – a new clone of *Plasmodium vivax* with high quality genome assembly ID 6525”; accession number ERP129582.

Illumina:

13=1a - ERS6867716
14=1b - ERS6867717
15=2a - ERS6867718
16=2b - ERS6867719
Pacbio:

DN599117N-A1 or 5987STDY8548200 - ERS3947829

### Genome assembly and annotation

PacBio subreads from sample DN599117N-A1 (5987STDY8548200) were used for the assembly. Circular consensus sequencing reads (CCS) were generated from the subreads using PacBio SMRTLink (https://www.pacb.com/support/software-downloads/).

Illumina reads: Samples 4472STDY7698313 (volunteer 1a), 4472STDY7698314 (volunteer 1b) and 4472STDY7698315 (volunteer 2a) from STDY4472 were used for assembly polishing. The Illumina reads were processed with CutAdapt 2.7 (72) to remove adapter sequences.

Decontamination of the sequencing data: To identify contaminant species in the sequencing data, BLAST searches of a randomly selected subset of subread sequences were run against the NCBI nt and nr databases (February 2020 versions) (73). The list of detected contaminants was then extended using Diamond 0.9.22 (74) BLASTX of the PacBio subreads and CCS reads against a database that contained 39,920 apicomplexan protein sequences and 190,075 protein sequences from various bacterial and fungal species. Further classification of sequences by species was performed using BLAST of the CCS reads and subreads against a nucleotide database containing the *P. vivax* P01 reference genome (30) (PlasmoDB release 46), GRCh38.p13 human genome assembly and 225 bacterial or fungal sequences. In parallel with this, the same reference nucleotide sequences were also used for competitive mapping of the PacBio and Illumina reads with Minimap2 2.17-r941 (75). Next, a Kraken 2.0.8-beta (76) database was made that contained multiple reference genomes of *Plasmodium*, as well as the genomes of human and other contaminant species that were detected in the previous steps. This database was used to classify the PacBio subreads, CCS reads and Illumina reads. If a PacBio subread was unambiguously detected as belonging to a contaminant species, other subreads that had been produced from the same zero-mode waveguide (ZMW) were also flagged as contaminants. Information from different contaminant detection methods was combined. BLAST against the NCBI nt database (October 2019 version) was run with the PacBio sequences that still remained unclassified after the previous steps. Illumina reads that remained unclassified after running Kraken were removed from the dataset.

In order to verify the effectiveness of Illumina read set decontamination, the Illumina reads were assembled with LightAssembler (initial public release version) (77), and the resulting assembly was checked for contaminants using Diamond BLASTX. The database for Diamond was the same as previously described for the decontamination of PacBio subreads. No contaminant sequences were detected in the Illumina assembly.

Canu assembly of PacBio data: Palindromic CCS were detected using a script from PacBio’s GitHub repository (https://github.com/PacificBiosciences/apps-scripts/blob/master/miscUTILS/missing_adaptors.py). The palindromic CCS were removed from the dataset. Next, the decontaminated PacBio subreads and CCS were pooled and assembled with Canu assembler (78) (with default settings of the pacbio-raw mode, and with the genomeSize=29052596 flag). The resulting assembly was deduplicated by merging contigs with unique overlaps using GAP5 v1.2.14-r3753M (79). The assembly was polished using the Arrow algorithm in PacBio gcpp (version 1, https://github.com/PacificBiosciences/gcpp), followed by three iterations of Pilon 1.23 (80). Decontaminated Illumina reads (pooled from three samples) were used as the input for Pilon. The apicoplast and mitochondrion sequences were circularized using Circlator minimus2 (81). Assembly completeness was assessed using BUSCO 3.0.1 (82). The assembly was annotated using the Glasgow server of Companion (83) (http://protozoacompanion.gla.ac.uk/, February 2020 version). The alignment of proteins to the reference genome was enabled in the Companion run and the rest of the settings were left as default.

Mapping of Illumina reads to estimate the coverage of specific genes: Pooled Illumina reads from three samples that had been processed with CutAdapt and decontaminated with Kraken were mapped to the assembly with Minimap2 using the short read mapping mode (“-ax sr”).

### VIR gene analysis

Analysis of the diversity and relatedness amongst the VIR genes of PvW1 compared to PvP01, PvT01, PvC01 and Sal-1 was carried out as described in (30).

### Statistical analysis

Unless otherwise stated, data were analyzed using GraphPad Prism version 9.1.1 for Windows (GraphPad Software Inc.). All tests used were 2-tailed and are described in the text. A value of *P*<0.05 was considered significant.

