## Supplemental Info for "Controlled human malaria infection with PvW1 – a new clone of *Plasmodium vivax* with high quality genome assembly"

### Supplementary Text

#### VAC068: Long-term safety monitoring

Follow-up clinic visits at dC+45 and dC+90 gave rise to no safety concerns or indication of relapsing infection, with negative qPCR readings recorded at both time-points. Ongoing annual follow-up by email will continue for 5 years post-CHMI, however, as of time of writing (May 2021; 3 years post-primaquine treatment) no relapse of *P. vivax* has been diagnosed for either volunteer. One volunteer did report symptoms of rapid-onset fever, chills, myalgia and headache, following a prodrome of fatigue and sore throat, 12 months post-CHMI. However, immediate testing for *P. vivax* by qPCR (plus RDT and thick film microscopy) was negative and fevers resolved with no specific treatment within 3 days, with residual symptoms fully resolving within 6 days. The working diagnosis was an intercurrent viral infection (blood tests showed a leukocytosis (11.96 x$10^9$/mL) with a relative grade 1 neutrophilia (10.7 x$10^9$/mL) and grade 3 lymphocytopenia (0.47 x$10^9$/mL); CRP 37 mg/L; urea, creatinine and liver function tests within normal limits). The same volunteer had a similar short-lived febrile illness 20 months and then 22 months post-CHMI but both resolved spontaneously after a few days with no specific treatment and no need for medical attendance.

### Supplementary Figures

**A**

| Mosquito Batch Number | Mosquitoes Dissected for Oocyst Count (n) | Mean [range] Oocyst Count on Days 6-7 | Mosquitoes Dissected for Spz Count (n) | Median [range] Spz Score in Salivary Glands on Day 14 | PvCSP-typing | Genotyping |
|---|---|---|---|---|---|---|
| **C01-001** | 5 | 9 [4-12] | N.D. | N.D. | VK210 | Probably mixed-infection |
| **C05-001** | 10 | 3 [0-6] | 10 | +2 [0 to +4] | VK210 | Mono-infection |
| **C05-002** | 5 | 19 [15-35] | N.D. | N.D. | VK210 | Mixed-infection (2 strains) |

**B**

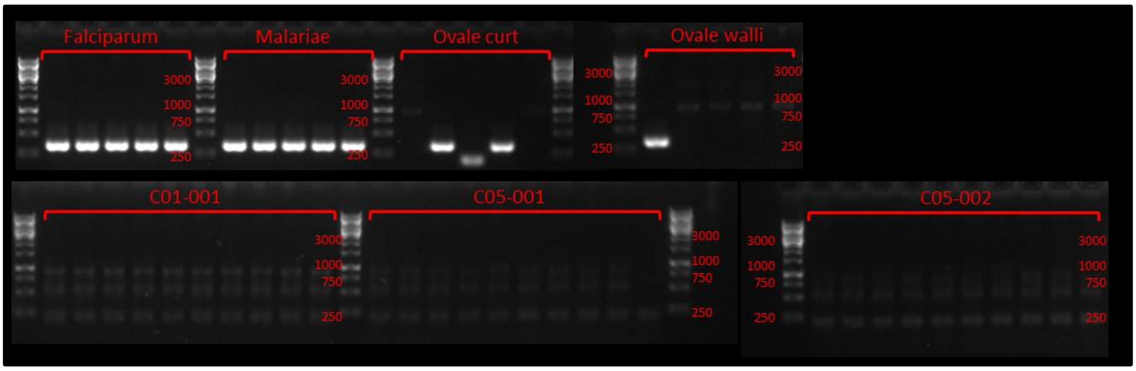


**C**

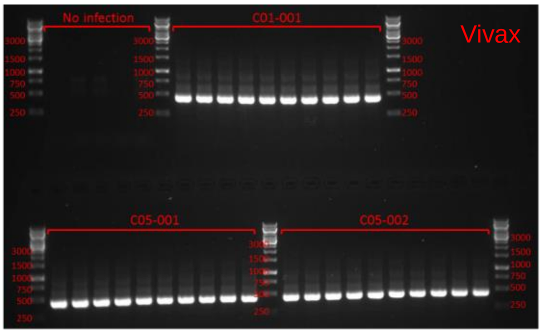


**Figure S1. Results of mosquito batch analyses conducted in real-time.**

(**A**) Three independent batches of mosquitoes were prepared in Thailand. Oocyst count data from dissected midguts and sporozoite (spz) count data from dissected salivary glands are reported. Score of +2 indicates >10-100 spz observed; score of +4 indicates >500 spz observed; 0 indicates negative; N.D. = not done. Genotyping of patient blood samples was performed to assess multiplicity of *P.*

*vivax* infection, with extracted DNA processed using a SNP barcode panel as described by the MalariaGEN network (1). Out of the three samples, the barcode sequences for two contained heterozygous genotype calls, suggesting the presence of more than one genotype within the infection. One blood sample (C05-001) had homozygous calls at all single nucleotide polymorphisms (SNPs), suggesting that it contained only a single *P. vivax* genotype. (**B**) Nested PCR to test for presence of *P. falciparum*, *P. malariae*, *P. ovale curtisi* and *P. ovale wallikeri* DNA. Amplified band at ~300 bp shown presence of DNA from each species of *Plasmodium* parasite. Positive control samples were tested in five replicates, with successful amplification in 5/5 cases for *P. falciparum* and *P. malariae*, but less reliable amplification for the two subspecies of *P. ovale*. Each of the three donor blood samples used to produce the batches of mosquitoes were tested in ten replicates. All test samples were negative. (**C**) The same PCR assay was performed using primers for *P. vivax*. All ten replicates for each donor blood sample were positive. Non-infected blood sample control was negative (tested in five replicates).

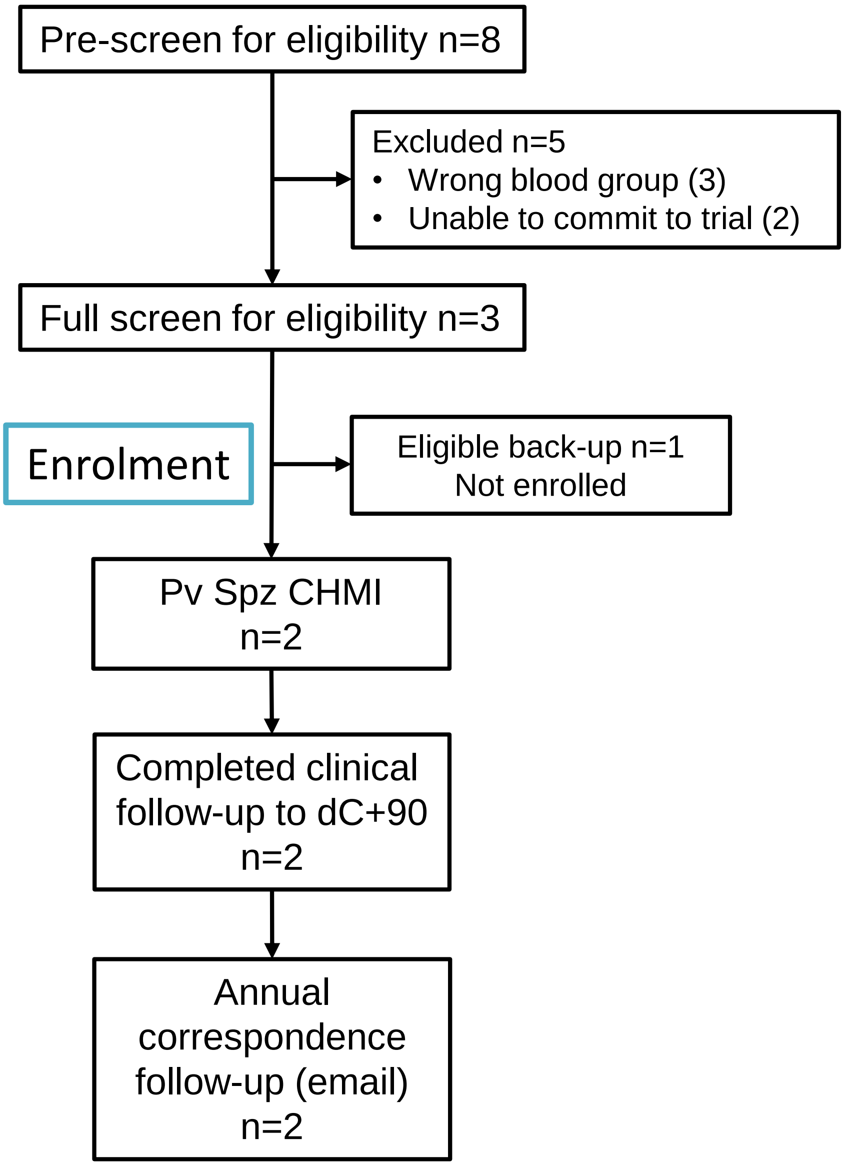


**Figure S2. VAC068 flow chart of study design and volunteer recruitment.**

Enrolment into the VAC068 study began in April 2018. Eight volunteers underwent pre-screening for O- blood group prior to full screening. Three eligible volunteers were identified, of whom one was a back-up (in case a volunteer withdrew prior to CHMI) and two proceeded to enrolment and underwent *P. vivax* CHMI delivered by mosquito-bite on 13 April 2018. Clinical follow-up continued until 90 days after challenge (dC+90) and was completed by 16 July 2019. Annual follow-up by email correspondence will continue for 5 years. Volunteer demographics are summarized in **Table S1**.

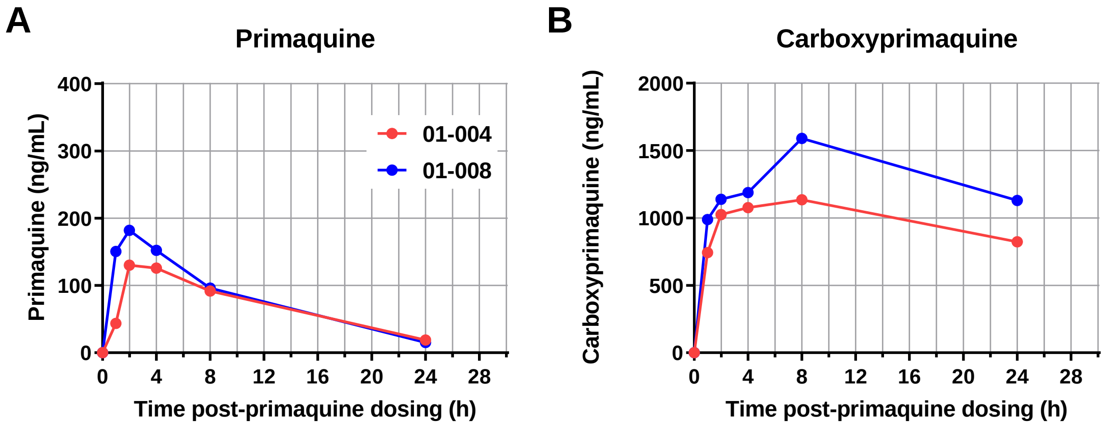


C

| Primaquine kinetic parameter | Vol ID: 01-004 | Vol ID: 01-008 |
|---|---|---|
| $C_{max}$ (ng/mL) | 130.3 | 181.8 |
| $t_{max}$ (h) | 2 | 2 |
| $AUC_{inf}$ (h*ng/mL)† | 1,878 | 2,094 |
| t½ (h)† | 7.2 | 6 |

**Figure S3. Results of primaquine test dose at screening for the VAC068 volunteers.**

Plasma concentrations of (**A**) primaquine (parent drug) and (**B**) carboxyprimaquine (metabolite) versus time in the two VAC068 volunteers following a single 30 mg oral dose of primaquine. Results for carboxyprimaquine, the major observed plasma metabolite of primaquine, are shown for completeness, but this is not produced primarily via CYP2D6 metabolism (2) and is, therefore, an unproven indicator of CYP2D6 phenotype and primaquine efficacy. (**C**) Pharmacokinetic parameters of primaquine. † Data previously reported by Bennett *et al.* (3) indicated that these kinetic parameters were significantly different between subjects that relapsed with *P. vivax* versus those that did not, consistent with decreased metabolism of primaquine. The primaquine datasets for both volunteers in VAC068 are consistent with the expected ranges for CYP2D6 extensive metabolizers as reported by Bennett *et al.* (3).

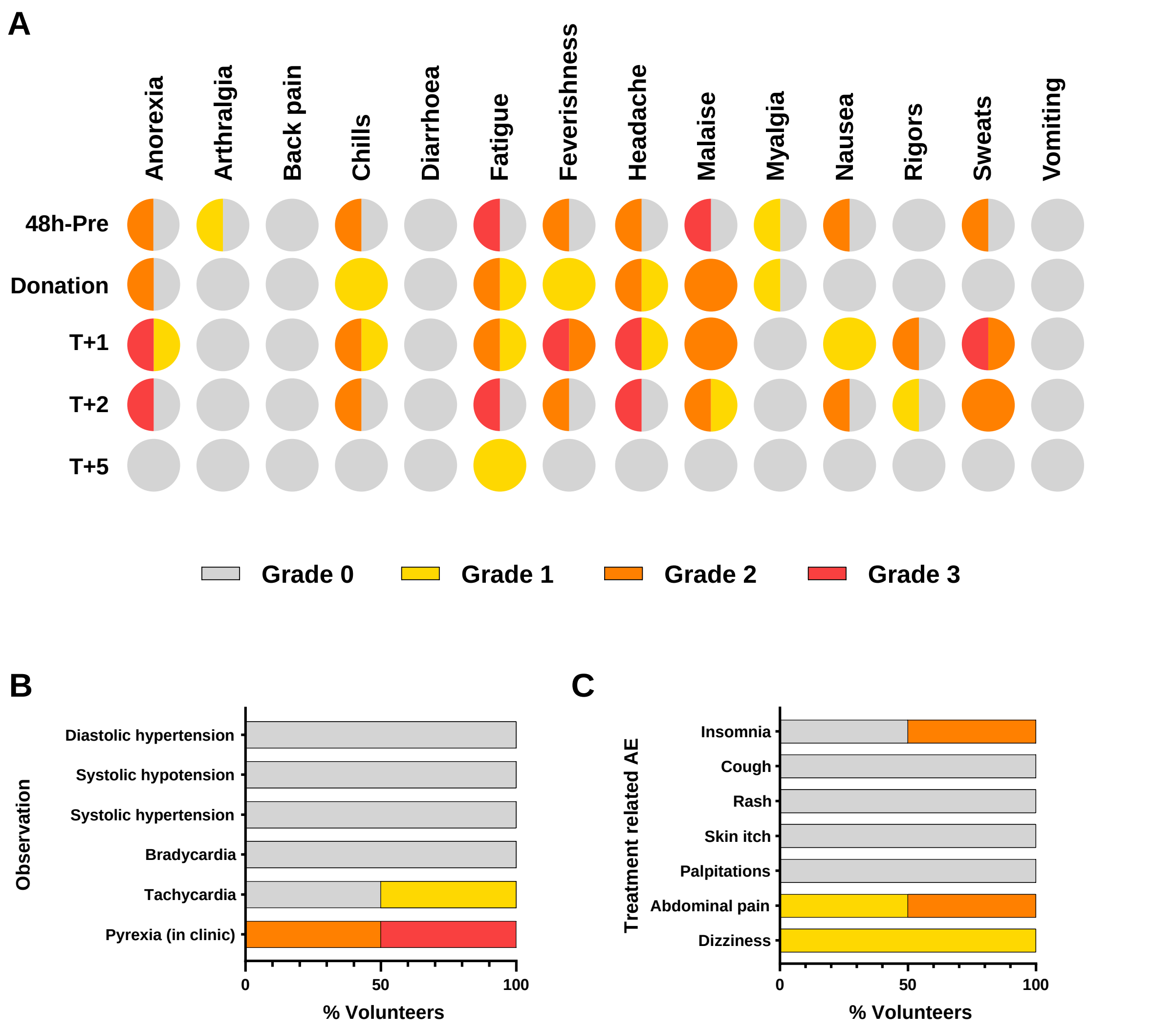


**Figure S4. VAC068 adverse event data following *P. vivax* sporozoite CHMI.**

(**A**) The solicited systemic AEs recorded at the indicated time-points during the CHMI period are shown as the maximum severity reported by each volunteer and as a percentage of the volunteers reporting each individual AE (n=2). Color-coding refers to AE grading: 0 = none; 1 = mild; 2 = moderate; 3 = severe. 48h-pre = the 48 hour period prior to blood donation; Donation = time-point of blood donation; +1, +2 and +5 days post-treatment (T). (**B**) Percentage of all volunteers (n=2) with AEs in recorded physical observations during the CHMI period (from day of challenge up until 90 days post-challenge). Pyrexia relates to the measured temperature during clinic visits. (**C**) AEs possibly relating to anti-malarial treatment (from day T+1 until completion of primaquine treatment course on T+16) shown as a percentage of all volunteers (n=2).

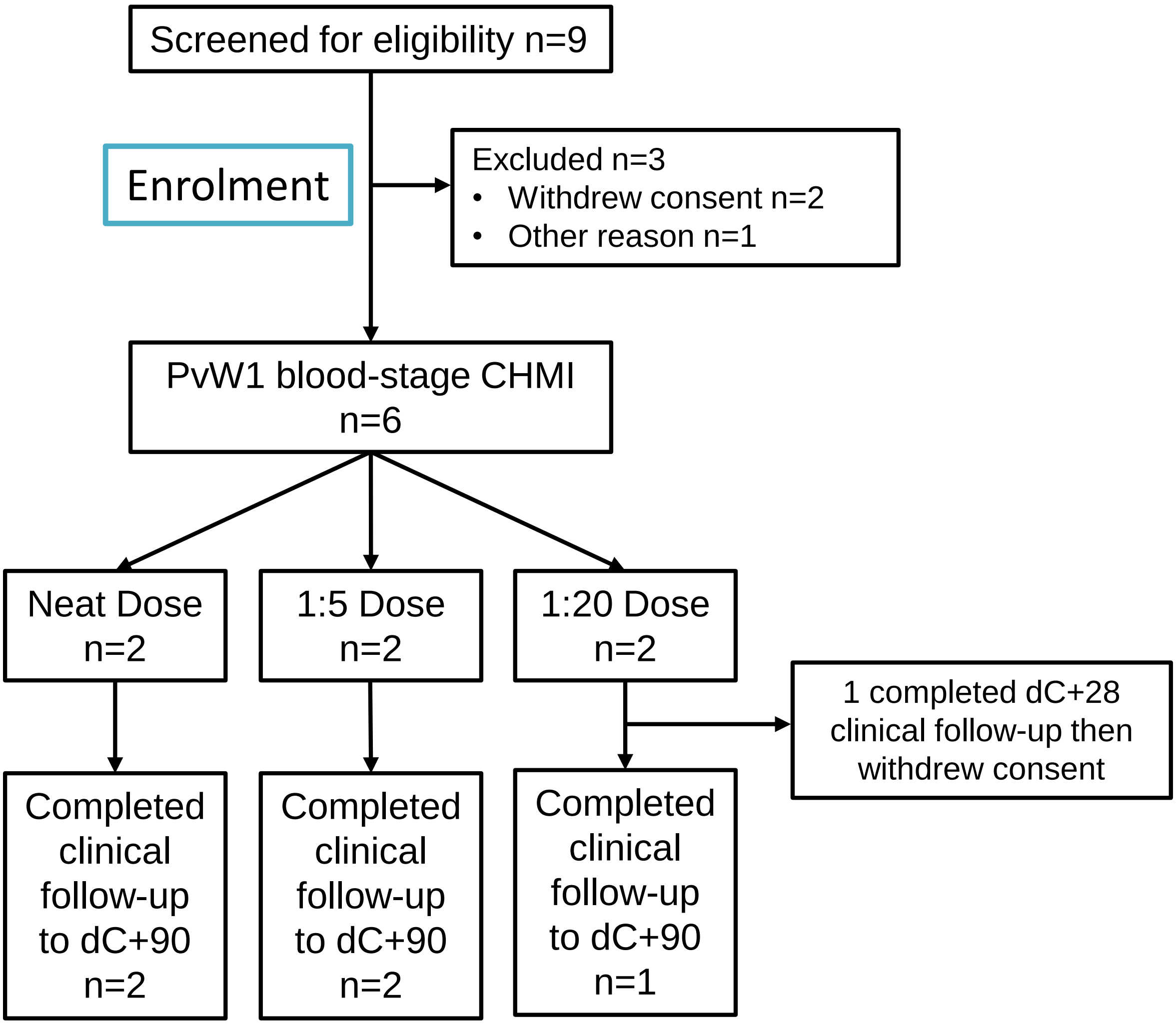


**Figure S5. VAC069A flow chart of study design and volunteer recruitment.**

Enrolment into the VAC069A study began in January 2019. Six volunteers (of the nine screened) were enrolled into the blood-stage CHMI trial – two into each dose group (neat, 1:5, 1:20 dilution). One volunteer in the 1:20 dose group completed the C+28 follow-up but then withdrew consent. The five remaining volunteers completed their follow-up until C+90. Across the groups, 4 males and 2 females were enrolled, the mean age was 27 years (range 18 – 33 years). None had previously travelled to malaria-endemic regions.

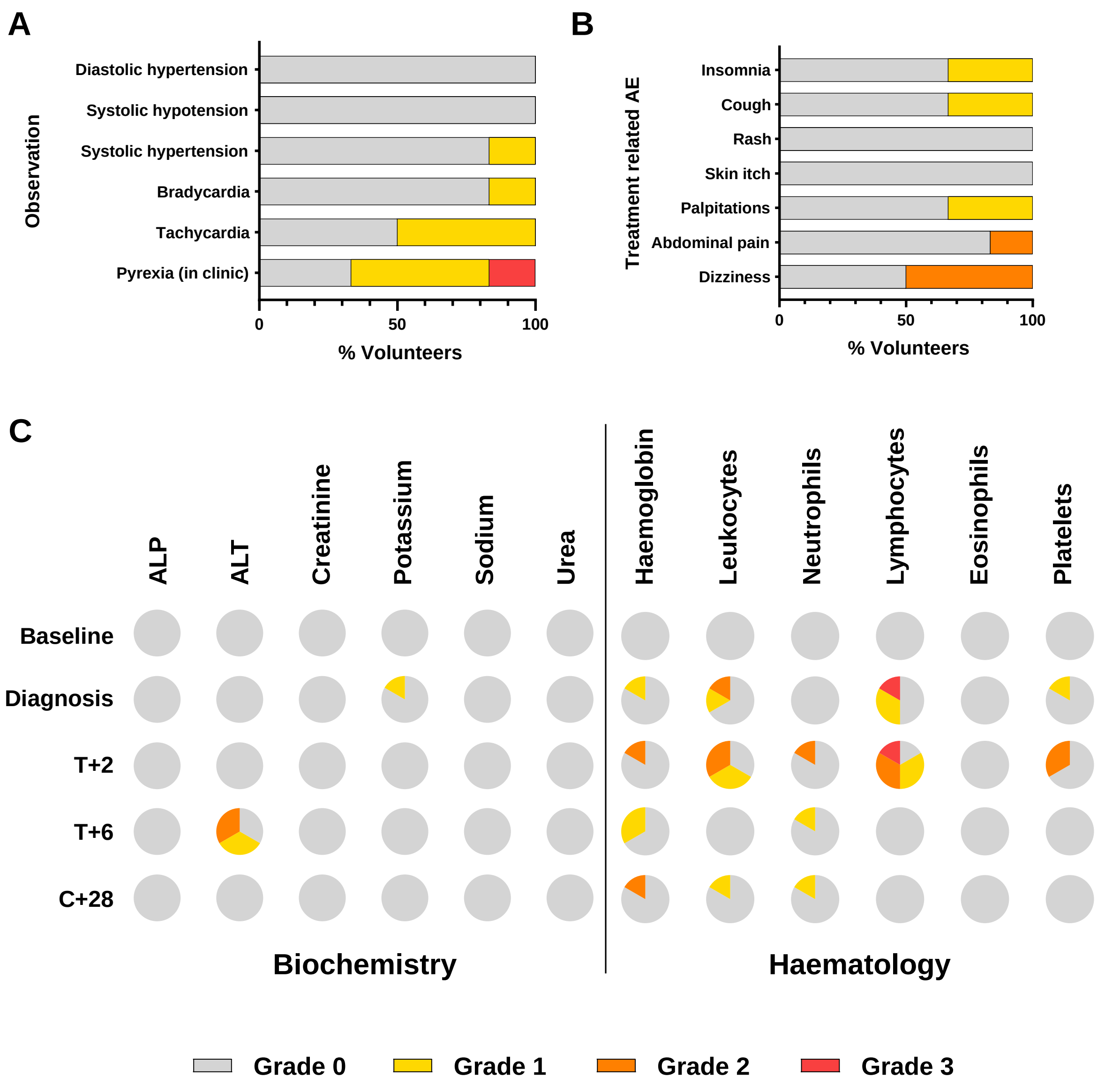


**Figure S6. VAC069A adverse event data following *P. vivax* PvW1 blood-stage CHMI.**

(**A**) Percentage of all volunteers (n=6) with AEs in recorded physical observations (from day of challenge up until 90 days post-challenge). Pyrexia relates to the measured temperature during clinic visits. (**B**) AEs possibly relating to anti-malarial treatment, recorded from day after starting treatment (T+1) until 28 days post-challenge (C+28, approximately 10 days post-completion of treatment), shown as a percentage of all volunteers (n=6). Color-coding refers to AE grading: 0 = none; 1 = mild; 2 = moderate; 3 = severe. **(C)** The laboratory AEs recorded at the indicated time-points during the CHMI period (day of challenge until 90 days post-challenge) are shown as the maximum severity reported for each volunteer and as a percentage of the volunteers reporting each individual AE (n=6). Baseline = prior to CHMI (C-1); Diagnosis = time-point of malaria diagnosis; +2 and +6 days post-treatment (T+2, T+6); C+28 = 28 days post-CHMI.

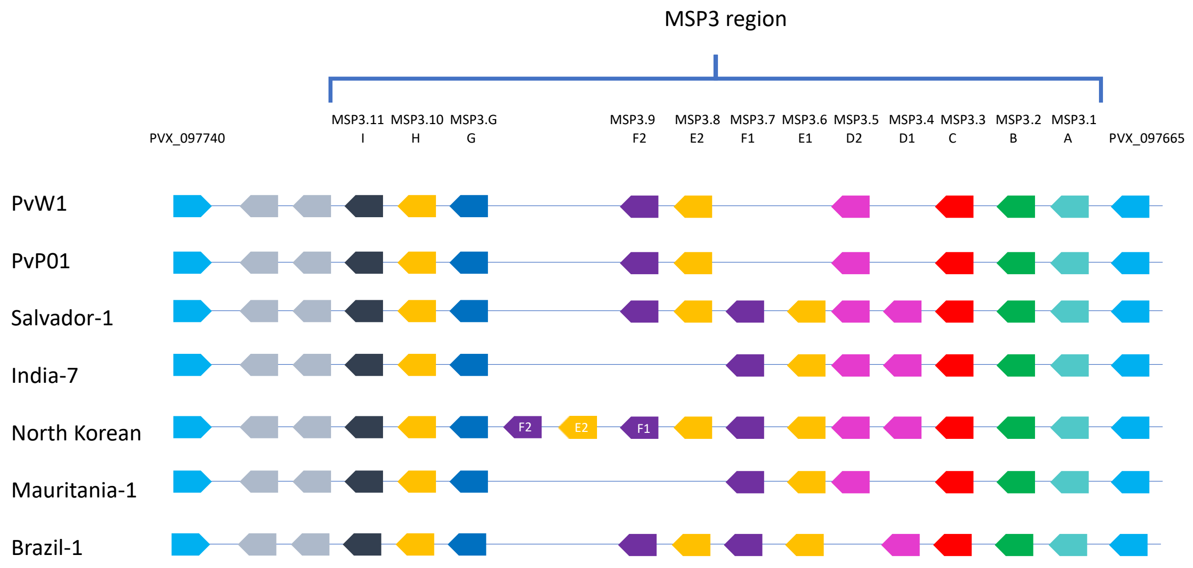


**Figure S7. Organization of the MSP3 multigene family in PvW1 compared to other *P. vivax* isolates.**

Organization of the MSP3 multigene family in PvW1 is compared to *P. vivax* isolates: PvP01 (4) and Salvador-1, India-7, North Korean, Mauritania-1, Brazil-1 (5). Flanking genes are shown and are syntenic across all isolates, as are MSP3.1, MSP3.2, MSP3.3, MSP3.G, MSP3.10 and MSP3.11. There is variability in the central region of the MSP3 region in the presence or absence and copy number of MSP3.4, MSP3.5, MSP3.6, MSP3.7, MSP3.8 and MSP3.9.

#### A PvCSP

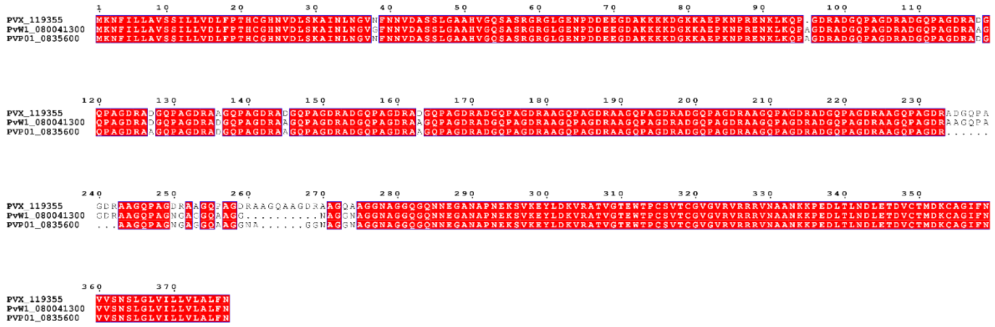


#### B Pvs25

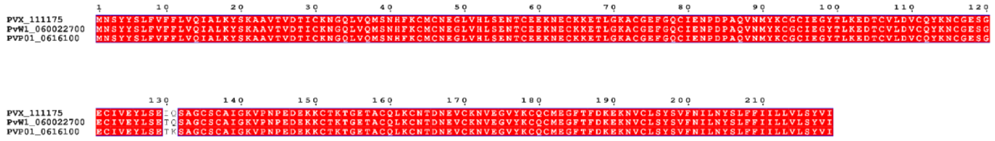


#### C PvDBP

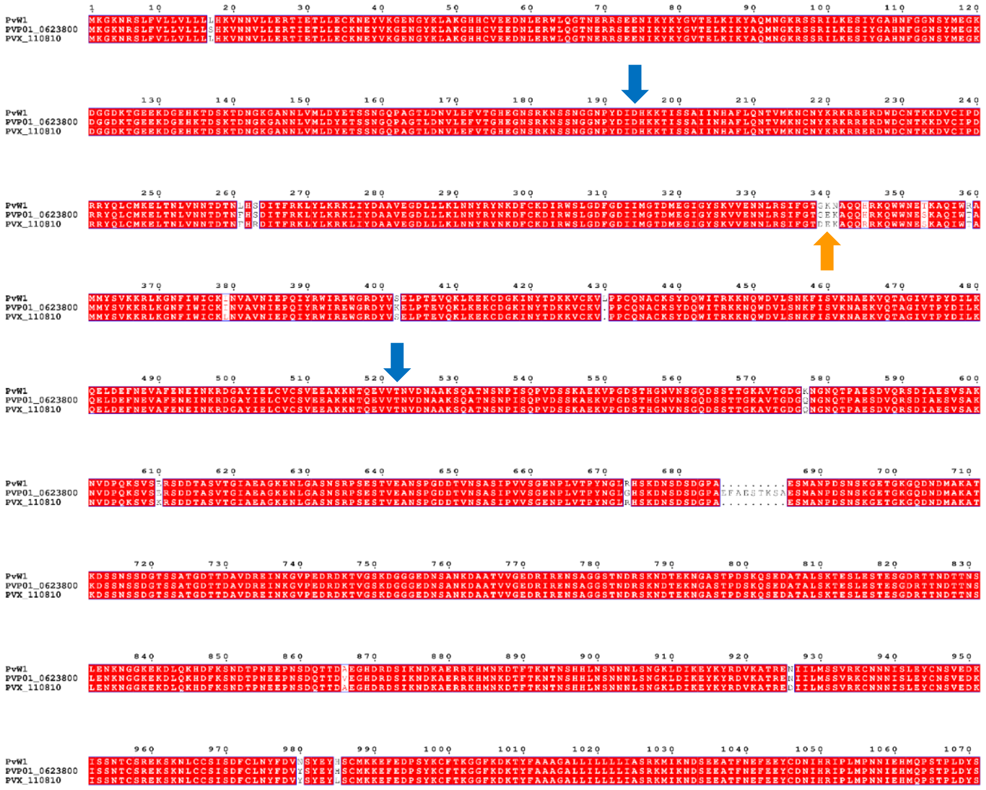

#### Figure S8. Amino acid sequence alignments of leading *P. vivax* vaccine candidate antigens.

Sequences are aligned for (**A**) sporozoite-stage PvCSP, (**B**) transmission-stage Pvs25, and (**C**) blood-stage PvDBP from PvW1, PvP01 and SalI (labelled PVX). The start and end of region II in PvDBP, present in two clinical vaccine candidates (6, 7), are indicated by the blue arrows (D194-T521, numbered as in SalI). The “DEK epitope” (8) is indicated by the orange arrow.

A

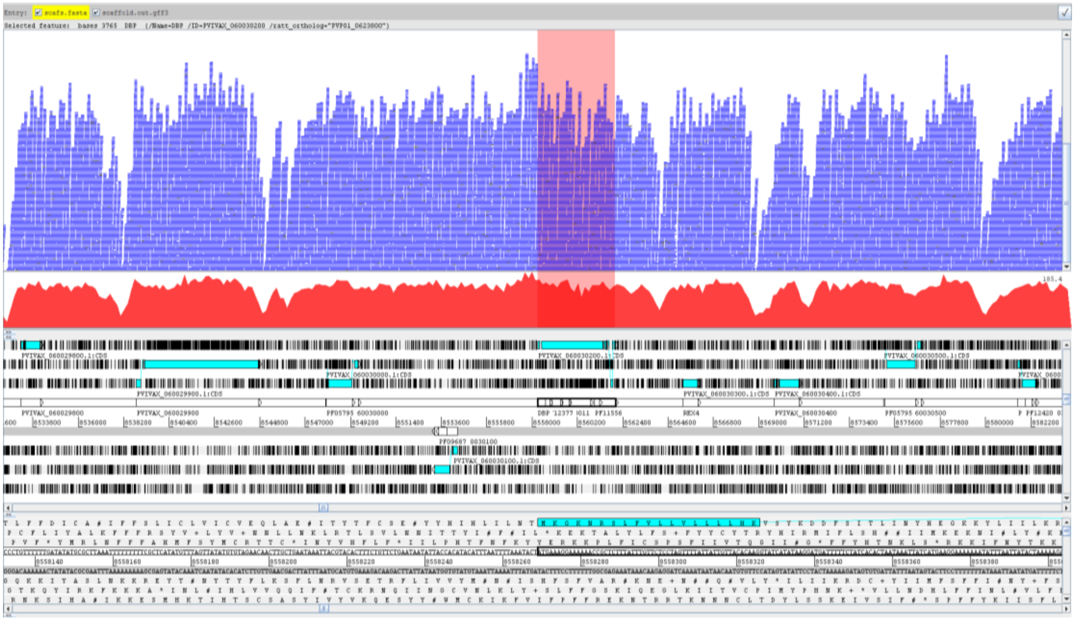

B

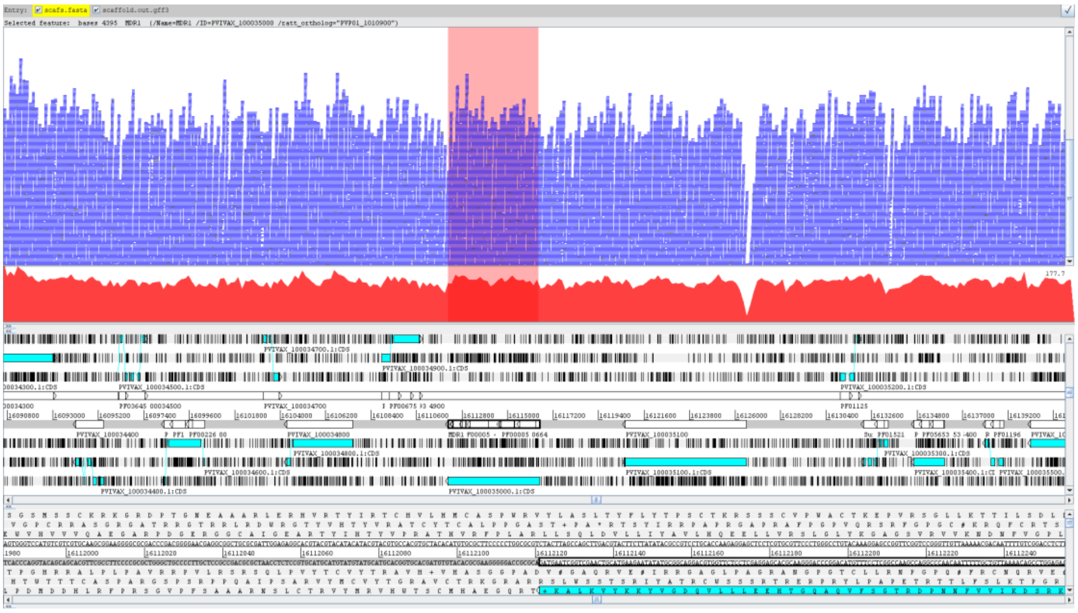

**C**

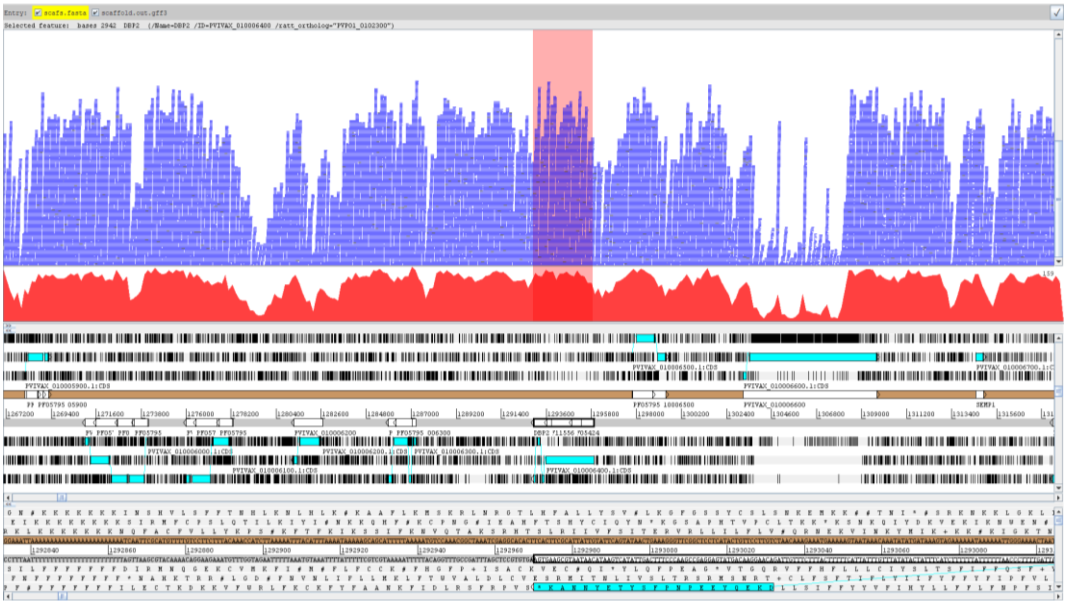

**D**

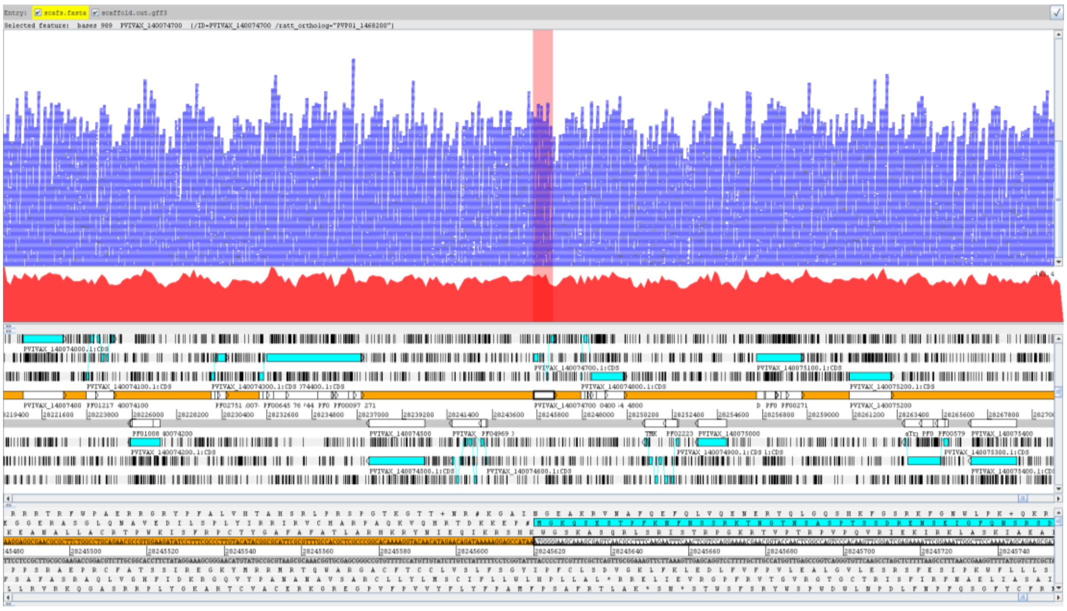

**Figure S9. Analysis of gene copy number variation in PvW1.**

Illumina reads were mapped to the new PvW1 genome assembly to determine the copy number of genes known to be present in multiple copies in some *P. vivax* isolates. Regions analyzed were (**A**) PvDBP; (**B**) PvMDR1; (**C**) PvDBP2; and (**D**) PvP01_1468200. Individual reads (blue) and coverage (red) are mapped above the genome assembly. The individual genes of interest are highlighted in pink. In all cases Illumina read coverage over the gene of interest appears to be consistent with the flanking regions suggesting that these genes are present in single copy in the PvW1 genome. Note that the genes labelled PVIVAX are in fact PvW1.

#### Supplementary Tables

| Demographic | Volunteer 1 | Volunteer 2 |
|---|---|---|
| **Sex** | Female | Male |
| **Age (5-year range)** | 30-35 | 20-25 |
| **BMI** | 19.8 | 23.2 |
| **Ethnicity** | Caucasian | Caucasian |
| **Blood group (ABO and rhesus)** | O negative | O negative |
| **Duffy blood group sero-phenotype** | Fya+Fyb+ | Fya+Fyb+ |
| **G6PD (U/gHb)[a]** | 9.0 | 7.7 |
| **HLA-type** | Typing Done | Typing Done |
| **Hemoglobinopathy screen** | Negative | Negative |
| **CMV sero-status** | Positive | Positive |
| **EBV sero-status** | Positive | Positive |
| **CYP2D6 genotype (predicted phenotype[b])** | CYP2D6*1/*1 (EXT) | CYP2D6*1/*4 (EXT) |
| **CYP2D6 metabolizer phenotype predicted by Activity Score (AS)-Model A[c]** | Score 2 (EXT) | Score 1 (EXT) |

**Table S1. Demographic information for the two volunteers in the VAC068 study.**

[a] Normal G6PD range = 5.8 – 18.8 U/gHb.

[b] Predicted CYP2D6 metabolizer phenotype according to (9) by service provider.

[c] Predicted CYP2D6 metabolizer phenotype according to (10), as also used in the study by Bennett *et al*. (3). AS-Model A scores range from 0 to 2, with 0 indicating poor or no CYP2D6 activity, 0.5 intermediate, and 1 or 2 indicating normal levels of CYP2D6 activity.

EXT = extensive metabolizer.

| Volunteer | DoD | D6.5 | D7 | D7.5 | D8 | D8.5 | D9 | D9.5 | D10 |
|---|---|---|---|---|---|---|---|---|---|
| 01-004 | 14.5 | N | N | N | 8 | 54 | 80 | 34 | 181 |
| 01-008 | 14 | N | N | N | N | 79 | 119 | 119 | 155 |

| Volunteer | DoD | D10.5 | D11 | D11.5 | D12 | D12.5 | D13 | D13.5 | D14 |
|---|---|---|---|---|---|---|---|---|---|
| 01-004 | 14.5 | 302 | 666 | 1036 | 1468 | 2517 | 8466 | 11449 | 18076 |
| 01-008 | 14 | 394 | 1026 | 1717 | 1865 | 2889 | 8841 | 16388 | 16717 |

| Volunteer | DoD | D14.5 | T+1 | T+2 | T+4 | T+10 | T+16 | C+45 | C+90 |
|---|---|---|---|---|---|---|---|---|---|
| 01-004 | 14.5 | 31010 | 610 | 0.2 | 1.1 | N | N | N | 0.4 |
| 01-008 | 14 |  | 52 | 0.9 | N | N | N | N | 0.0 |

**Table S2: Raw qPCR data (genome copies/mL) for VAC068.**

Top row represents day of follow-up visit post mosquito-bite CHMI. Data highlighted in red represent qPCR measurement at time-point of blood donation, also referred to as "day of donation" (DoD) for a particular individual. N = PCR negative for all three triplicate readings in the assay. Squares highlighted in grey indicate negative or a mean reading <20 gc/mL (below minimum positive reporting criteria). Squares highlighted in blue indicate samples taken post-treatment: T+1 indicates 1 day immediately post-treatment initiation; longer-term clinical follow-up occurred at days C+45 and C+90.

**A**

| MEDDRA System Organ Class | MEDDRA Higher Level Term | MEDDRA Preferred Term | Subject ID (diagnosis day) | Onset (days post-CHMI) | AE duration (days) | Maximum severity |
|---|---|---|---|---|---|---|
| Gastrointestinal disorders | Gastrointestinal and abdominal pains (excl oral and throat) | Abdominal pain upper | 01-004 | 11 | 2 | Grade 1 |
| Musculoskeletal and connective tissue disorders | Joint related signs and symptoms | Arthralgia | 01-004 | 12 | 2 | Grade 1 |
| Musculoskeletal and connective tissue disorders | Musculoskeletal and connective tissue pain and discomfort | Back pain | 01-004 | 13 | 1 | Grade 1 |
| Cardiac disorders | Cardiac signs and symptoms NEC | Dizziness | 01-008 | 11 | 1 | Grade 1 |

**B**

| MEDDRA System Organ Class | MEDDRA Higher Level Term | MEDDRA Preferred Term | Subject ID | Onset (days post-CHMI) | AE duration (days) | Maximum severity |
|---|---|---|---|---|---|---|
| Nervous system disorders | Migraine headaches | Migraine | 01-004 | 66 | 2 | Grade 3 |

**C**

| Laboratory AE | Volunteer | Time-point | Result | Grade | Action taken |
|---|---|---|---|---|---|
| Hyber-bilirubinaemia (µmol/L) | 01-008 | C-1 | 32 | 2 | No clinical concern, normal ALT/ALP. Resolved by C+14 (20 µmol/L). No action taken. |
| Anaemia (g/L) | 01-008 | C+47 | 123 | 1 | No clinical concern, resolved by C+94 (13.1 g/L). No action taken. |
| Low white blood cell count ($x10^9$/L) | 01-004 | C+14 | 3.03 | 1 | Consistent with *P. vivax* diagnosis, repeated and resolved by C+46 (5.77 $x10^9$/L). |
| Lymphocyto-paenia ($x10^9$/L) | 01-008 | C+14 | 0.67 | 2 | Consistent with *P. vivax* diagnosis, repeated and resolved by C+47 (1.95 $x10^9$/L). |
| | 01-004 | C+14 | 0.56 | 2 | Consistent with *P. vivax* diagnosis, repeated and resolved by C+47 (1.89 $x10^9$/L). |

**Table S3: VAC068 unsolicited, grade 3 and laboratory AEs.**

(**A**) Unsolicited AEs deemed at least possibly (possibly, probably or definitely) related to *P. vivax* sporozoite CHMI in all volunteers (n=2) are shown, with MedDRA coding and maximum severity reported. (**B**) Onset and duration of grade 3 AEs: only one grade 3 unsolicited AE (not related to CHMI) was reported by volunteer 01-004. Headache, starting at C+66, lasted 2 days and involved attendance to the volunteer's medical practitioner, but resolved fully. The reported headache was very similar to the volunteer's previous migraines. Grade 3 solicited AEs are shown in **Fig. 1B** and **S4**. (**C**) Laboratory AEs in all volunteers (n=2) in the VAC068 study, from day of challenge until 90 days post-challenge. Blood was drawn for haematology and biochemistry pre-CHMI (day C-1) and post-CHMI on days C+9, C+11, on day of blood donation (C+14) with an additional draw for haematology within 12 hours of treatment, and finally at C+45 and C+90. Blood tests were also carried out at other time-points if clinically indicated. Maximum reported severity is shown, graded as per site-specific grading tables; (Grading: 1 = mild; 2 = moderate; 3 = severe). ALT = alanine aminotransferase; ALP = alkaline phosphatase.

| Volunteer | DoD | D0 | D1 | D2 | D2.5 | D3 | D3.5 | D4 | D4.5 | D5 | D5.5 | D6 | D6.5 |
|---|---|---|---|---|---|---|---|---|---|---|---|---|---|
| **01-002** | **15.5** | N | N | N | N | N | N | N | N | N | 20 | N | 7 |
| **01-003** | **12.5** | N | N | N | N | N | N | N | N | 11 | 6 | 11 | 63 |
| **01-005** | **15** | N | N | N | N | N | N | N | N | N | N | N | N |
| **01-006** | **15** | N | N | N | N | N | N | N | N | N | N | N | N |
| **01-007** | **15.5** | N | N | N | N | N | N | N | N | N | N | N | N |
| **01-009** | **16.5** | N | N | N | N | N | N | N | N | N | N | 2 | 15 |

| Volunteer | DoD | D7 | D7.5 | D8 | D8.5 | D9 | D9.5 | D10 | D10.5 | D11 | D11.5 | D12 | D12.5 |
|---|---|---|---|---|---|---|---|---|---|---|---|---|---|
| **01-002** | **15.5** | 17 | 6 | 17 | 51 | 58 | 95 | 67 | 156 | 301 | 290 | 294 | 411 |
| **01-003** | **12.5** | 78 | 45 | 60 | 260 | 508 | 407 | 1060 | 2060 | 3132 | 2640 | 5393 | 11259 |
| **01-005** | **15** | N | 5 | 18 | 22 | 72 | 44 | 239 | 250 | 519 | 611 | 972 | 1735 |
| **01-006** | **15** | 6 | N | 21 | 21 | 33 | 38 | 8 | 227 | 266 | 317 | 459 | 986 |
| **01-007** | **15.5** | N | N | N | 23 | 32 | 58 | 57 | 204 | 406 | 207 | 616 | 1227 |
| **01-009** | **16.5** | N | N | N | 13 | 15 | 16 | 8 | 55 | 53 | 24 | 76 | 305 |

| Volunteer | DoD | D13 | D13.5 | D14 | D14.5 | D15 | D15.5 | D16 | D16.5 | D17 | D17.5 | D18 | D19 |
|---|---|---|---|---|---|---|---|---|---|---|---|---|---|
| **01-002** | **15.5** | 956 | | 2251 | 417 | 5035 | 3779 | | 182 | | 168 | | |
| **01-003** | **12.5** | | 17 | | N | | | | | | | N | |
| **01-005** | **15** | 2666 | 2999 | 4700 | 1538 | 17795 | 14843 | | 13925 | | 1324 | 246 | |
| **01-006** | **15** | 1331 | | 2632 | 1056 | 8687 | 7112 | | 294 | | 154 | | |
| **01-007** | **15.5** | 1431 | 1253 | 3054 | 1444 | 9597 | 8134 | 15768 | | 555 | | 355 | |
| **01-009** | **16.5** | 239 | | 717 | | 1479 | 1438 | 2593 | 9668 | 7894 | | 534 | 464 |

**Table S4: Raw qPCR data (genome copies/mL) for VAC069A.**

Top row represents day of follow-up visit post blood-stage CHMI. Data highlighted in red represent qPCR measurement at time-point of diagnosis according to protocol, also referred to as "day of diagnosis" (DoD) for a particular individual. N = PCR negative for all three triplicate readings in the assay. Squares highlighted in grey indicate negative or a mean reading <20 gc/mL (below minimum positive reporting criteria). Squares highlighted in orange indicate a sample taken immediately pre-treatment at the next clinic visit following diagnosis; where this occurs, volunteers were officially diagnosed based on symptoms and/or qPCR and/or thick film microscopy data obtained in real time between clinic visits. Squares highlighted in blue indicate samples taken post-treatment.

## A

| Volunteer | Grade 3 AE | Time-point | Persisted at Grade 3 | Resolution |
|---|---|---|---|---|
| 01-002 | Feverishness | T+1 | 24 hours | T+2 |
| 01-003 | Feverishness | Diagnosis/C+12.5 | 48 hours | T+2 |
| | Chills | T+1 | 24 hours | T+2 |
| | Sweats | T+1 | 24 hours | T+2 |
| | Myalgia | T+1 | 24 hours | T+2 |
| | Fatigue | T+1 | 24 hours | C+28 |
| | Malaise | T+1 | 24 hours | T+2 |
| 01-006 | Feverishness | T+1 | 24 hours | T+2 |
| | Chills | T+1 | 24 hours | T+2 |
| | Headache | T+1 | 24 hours | T+2 |
| | Fatigue | T+1 | 24 hours | T+2 |
| | Malaise | T+1 | 24 hours | T+2 |
| 01-009 | Feverishness | T+1 | 24 hours | T+2 |
| | Rigor | T+1 | 24 hours | T+2 |

## B

| MEDDRA System Organ Class | MEDDRA Higher Level Term | MEDDRA Preferred Term | Subject ID (diagnosis day) | Onset (days post-CHMI) | AE duration (days) | Maximum severity |
|---|---|---|---|---|---|---|
| Skin and subcutaneous tissue disorders | Pruritus NEC | Pruritus | 01-003 (C+12.5) | 0 | 1 | Grade 1 |
| Metabolism and nutrition disorders | Appetite disorders | Decreased appetite | 01-003 (C+12.5) | 11 | 4 | Grade 2 |
| Musculoskeletal and connective tissue disorders | Muscle pains | Myalgia | 01-006 (C+15) | 11 | 1 | Grade 1 |

## C

| Laboratory AE | Volunteer | Time-point | Result | Grade | Action taken |
|---|---|---|---|---|---|
| Anaemia (g/L) | 01-002 | T+6 | 119 | 1 | Repeated and resolved at next visit (C+28) |
| | 01-005 | C+16, C+34 (C+14-C+90) | 98 (98-109) | (1-)2 | Monitored from C+14 to C+90, AE ongoing at C+90 (102 g/L; Grade 1) Referred to General Practitioner for ongoing medical care, with follow-up of AE ongoing |
| Low white blood cell count ($x10^9$/L) | 01-002 | T+1 | 3.3 | 1 | Consistent with *P. vivax* diagnosis, repeated and resolved (6.53 $x10^9$/L) at next visit (T+6) |
| | 01-003 | T+1 | 2.93 | 1 | Consistent with *P. vivax* diagnosis, repeated and resolved (6.86 $x10^9$/L) at next visit (T+6) |
| | 01-005 | T+1 (C+7-C+90) | 1.68 (1.68-3.39) | (1-)2 | Monitored from C+7 to C+90, AE ongoing at C+90 (3.36 $x10^9$/L; Grade1) Referred to General Practitioner for monitoring / ongoing care |
| | 01-006 | T+1 (C+14-T+1) | 2.36 | 2 | Monitored from C+14 to T+6, resolved at T+6 (4.84 $x10^9$/L); consistent with *P. vivax* diagnosis |
| Thrombocyto-paenia ($x10^9$/L) | 01-003 | T+1 | 114 | 2 | Consistent with *P. vivax* diagnosis; repeated and resolved (233 $x10^9$/L) at next visit (T+6) |
| | 01-005 | T+1 (Diagnosis-T+1) | 115 (115-127) | (1-)2 | Consistent with *P. vivax* diagnosis; repeated and resolved at T+6 (219 $x10^9$/L) |
| Neutropaenia ($x10^9$/L) | 01-005 | C+45 (C+14-C+90) | 0.76 (0.76-1.46) | (1-)2 | Monitored from C+14 to C+90, AE ongoing at C+90 (1.26 $x10^9$/L; Grade 1) Referred to General Practitioner for monitoring / ongoing care |
| Lymphocyto-paenia ($x10^9$/L) | 01-002 | T+1 (Diagnosis-T+1) | 0.72 (0.72-0.86) | (1-)2 | Consistent with *P. vivax* diagnosis, repeated and resolved at T+6 (1.63 $x10^9$/L) |
| | 01-003 | T+1 | 0.77 | 1 | Consistent with *P. vivax* diagnosis, repeated and resolved at T+6 (2.53 $x10^9$/L) |
| | 01-005 | Diagnosis (Diagnosis-T+1) | 0.44 (0.44-0.63) | (2-)3 | Consistent with *P. vivax* diagnosis, repeated and resolved at T+6 (1.63 $x10^9$/L) |
| | 01-006 | T+1 (C+14-T+1) | 0.3 (0.3-0.83) | (1-)3 | Consistent with *P. vivax* diagnosis, repeated and resolved at T+6 (2.07 $x10^9$/L) |

## D

| Laboratory AE | Volunteer | Time-point | Result | Grade | Action taken |
|---|---|---|---|---|---|
| Hyponatraemia (mmol/L) | 01-003 | C+28 | 134 | 1 | No clinical concern, no action taken |
| | 01-003 | C+90 | 134 | 1 | No clinical concern, no action taken |
| Hypokalaemia (mmol/L) | 01-002 | C+90 | 3.3 | 1 | No clinical concern, no action taken |
| | 01-005 | Diagnosis | 3.3 | 1 | Repeated within 24 hours at next visit, resolved (3.7 mmol/L) |
| | 01-003 | C+90 | 3.3 | 1 | No clinical concern, no action taken |
| ALT (IU/L) | 01-003 | T+6 | 135 | 2 | Asymptomatic, repeated in 48 hours (80 IU/L; Grade 1) and then resolved at C+28 (21 IU/L) |
| | 01-006 | T+6 | 56 | 1 | Repeated at next visit (C+28) and resolved (29 IU/L) |
| | 01-007 | T+6 | 80 | 1 | Repeated at next visit (C+28) and resolved (31 IU/L) |
| | 01-009 | T+6 | 114 | 2 | Asymptomatic, repeated in 72 hours and resolved (40 IU/L) |

#### Table S5: VAC069A grade 3, unsolicited and laboratory AEs.

(**A**) Onset and duration of grade 3 solicited AEs reported by all volunteers (n=6) in the VAC069A study. (**B**) Unsolicited AEs deemed at least possibly (possibly, probably or definitely) related to *P. vivax* PvW1 blood-stage CHMI in all volunteers (n=6) in the VAC069A study. These were assigned a MedDRA code, with maximum severity reported. (**C**) Laboratory AEs in full blood count in all volunteers (n=6) in the VAC069A study. (**D**) Laboratory AEs in biochemistry profile (urea and electrolytes, liver function tests). ALT = alanine aminotransferase. Maximum reported severity is shown, graded as per site-specific grading tables; (Grading: 1 = mild; 2 = moderate; 3 = severe). In all panels, time-point(s) = days in relation to day of CHMI (C) or treatment (T).
